# Language attrition and semi-lingualism among Liberian and Sierra Leonean refugee children: A sociolinguistic-psychological dynamic of trauma and mental health in stateless refugees in Oru, Nigeria

**DOI:** 10.64898/2026.03.17.26348612

**Authors:** Dogbahgen Alphonso Yarseah, Olu Francis Ibimiluyi, Omowumi Omojola Awosusi, J. Mac-Nixon Flomo, Bello Folaranmi Fatai, Elijah Olawale Olaoye, Alade Folasade Adesola, Ololade Omolayo Ogunsanmi

**Affiliations:** Ekiti State University, Faculty of Education, Department of Guidance and Counseling; Ekiti State University, Faculty of Education, Department of Guidance and Counseling Ado Ekiti, Nigeria; Federal University of Oye-Ekiti Office of the VC Ekiti State, Nigeria; University of Liberia Amos C. Sawyer College of Social Sciences and Humanity Department of Sociology and Anthropology & Criminology; Cavendish University of Uganda, Department of International Relations; Ekiti State University, Faculty Arts, Department of French Ekiti State, Nigeria; Ekiti State University Facuty of Education, Department of Guidance and Counseling Ado Ekiti; Department of Public Health Babcock University, Ogun State, Nigeria

**Keywords:** Language attrition, bilingual competence, trauma exposure, refugee children, CPTSD, PTSD, functional impairment

## Abstract

**Background:** Liberian and Sierra Leonean children born during and after the 2012 UNHCR cessation clause, and the subsequent closure of the Oru refugee camp in Nigeria, have grown up in conditions of protracted displacement and de facto statelessness. Many of these children have been exposed to multiple forms of trauma, including witnessing violence as well as physical, emotional, and sexual adversities within a complex and resource-constrained environment. Many also experience cultural-linguistic disruptions, including heritage-language attrition and increased reliance on host-country languages, which may be associated with challenges in identity formation and social integration. However, little is known about how trauma exposure interacts with language-related factors to influence PTSD and complex PTSD (CPTSD)-related functional impairment among stateless refugee children.

**Methods:** Using a cross-sectional design, 320 children aged 6-17 years (180 Liberian, 140 Sierra Leonean) were assessed. Trauma exposure was measured using the Child and Adolescent Trauma Screen (CATS), and PTSD/CPTSD functional impairment using the International Trauma Questionnaire-Child and Adolescent Version (ITQ-CA). Heritage- and host-language proficiency were assessed using a structured sociolinguistic questionnaire. Multivariate covariance analyses were conducted using SPSS to examine main and interaction effects.

**Results:** Multivariate analyses revealed that poorer host-language communication was associated with higher PTSD-related functional impairment (F(3, 311) = 2.85, p = .038, partial eta-squared = .027), whereas CPTSD impairment was largely unaffected. Native-language proficiency also predicted PTSD impairment (F(3, 290) = 3.44, p = .017, partial eta-squared = .034), and children with low heritage-language skills, limited parental/home-language exposure, and no Nigerian-language use showed the highest CPTSD impairment. Emotional connection to the native language provided a modest protective effect. The combined heritage- and host-language exposure was linked to lower trauma-related functional impairment, particularly for children at higher risk of CPTSD. Witnessed trauma emerged as the strongest predictor of functional impairment among refugee children, with CPTSD outcomes showing greater sensitivity (partial eta-squared = .153) than PTSD (partial eta-squared = .076).

**Conclusions:** Heritage-language competence and bilingual proficiency were associated with reduced PTSD-related functional impairment, whereas CPTSD was more strongly shaped by cumulative relational trauma. These findings highlight the potential value of interventions that support bilingual development and heritage-language preservation as pathways to resilience among stateless refugee children.

## INTRODUCTION

### Background of the Study

Liberian and Sierra Leonean refugees have experienced prolonged exposure to armed conflict, displacement, and social instability since the early 1990s. Civil wars in Liberia (1989–2003) and Sierra Leone (1991–2002) were characterized by mass atrocities and widespread displacement, forcing hundreds of thousands into exile across West Africa. Nigeria received significant numbers of refugees from both countries (Osunkoya, 2025; USCR, 2001), many of whom remained in protracted displacement for decades, particularly in and around the former Oru Refugee Camp. Although the wars formally ended over two decades ago, many refugee families have been unable or unwilling to return, resulting in a generation of children born and raised in exile in statelessness.

The persistence of displacement among Liberian and Sierra Leonean refugees cannot be understood solely in terms of past violence but must be situated within postwar political and structural conditions. In Liberia, unresolved political and ethnic divisions and postwar governance challenges contributed to ongoing fears of exclusion, while in Sierra Leone, persistent poverty, weak governance, and social inequality have constrained sustainable reintegration (SDG Reports, 2024; 2025). These structural deficits have direct consequences for children, including limited access to education, healthcare, and social protection, highlighting ongoing challenges to SDG 4 (Quality Education), SDG 10 (Reduced Inequalities), and SDG 16 (Peace, Justice, and Strong Institutions). Within Kunz’s refugee typology, many of these families can be understood as self-alienated refugees (Kunz, 1981), whose psychological wounds and structural marginalization inhibit durable return, with children inheriting disrupted identities and fractured senses of belonging.

Socio-ecological research from Sierra Leone shows that emotional distress is shaped primarily by community- and societal-level stressors, including poverty, limited livelihoods, and weak infrastructure (Horn et al., 2021). Comparable structural conditions characterize former refugee camp settings such as Oru in Nigeria, where camp closure occurred without sustained integration or social protection, resulting in prolonged residence under conditions of precarity.

Following the UNHCR cessation clause in 2012, Liberian and Sierra Leonean refugees in Nigeria lost formal protections and entered legal ambiguity (Durodola, 2021). Children raised before, during, and after this period developed under overlapping adversities—housing insecurity, poverty, disrupted schooling, and repeated exposure to community and interpersonal trauma—that extended beyond discrete war-related events. Eligibility for resettlement under the 1951 Convention Relating to the Status of Refugees requires demonstrable evidence of a “well-founded fear of being persecuted for reasons of race, religion, nationality, membership of a particular social group, or political opinion” (UN General Assembly, 1951, Art. 1A[2]). UNHCR operationalizes these criteria through vulnerability categories, including survivors of torture or violence, women and girls at risk, individuals with urgent medical needs, and persons facing serious protection threats (UNHCR, 2011; 2021). Many refugee parents, however, lacked literacy and knowledge of international law, framing claims around personal experiences of trauma, loss, and economic hardship. Although deeply distressing, these claims often fell outside formal legal thresholds, leading to repeated failures and extended uncertainty.

This gap between legal requirements and parental understanding had severe consequences. With resettlement denied and repatriation to Liberia or Sierra Leone unfeasible, over 40 family heads were left without durable legal status. Restrictive naturalization and integration policies in Nigeria further blocked access to citizenship, leaving families in prolonged legal limbo and de facto statelessness. For children growing up under these conditions, statelessness functioned not merely as a legal classification but as a structural and ecological environment shaping schooling, family interaction, identity formation, and intergenerational language transmission.

Within forced displacement contexts, language represents a frequently overlooked yet structurally consequential dimension of children’s psychosocial development. Stateless refugee communities in Nigeria are linguistically heterogeneous, encompassing Liberian English varieties, Sierra Leonean Krio, multiple indigenous languages, and host-community languages such as Yoruba or Igbo. Disrupted schooling, economic precarity, and weakened community cohesion place the burden of language transmission primarily on the home, where parents’ own linguistic marginalization constrains intergenerational continuity, resulting in unstable and fragmented linguistic repertoires among children.

Historically, such patterns were interpreted through deficit-oriented frameworks of semi-lingualism, most notably in Hansegård’s (1968) claim that incomplete mastery of the mother tongue reflected a “deviant psyche” and produced impoverished emotional and cognitive life. However, subsequent scholarship has decisively challenged this pathologizing view, demonstrating that fragmented language development is better understood as an outcome of structural inequality, interrupted education, and institutional language suppression rather than intrinsic psychological deficit (Skutnabb-Kangas, 1984; Karlander & Salö, 2026).

For Liberian refugees, these dynamics are rooted in long-standing historical language hierarchies: since independence, Liberia has maintained an English-only system of governance and education despite the presence of at least sixteen largely mutually unintelligible indigenous languages, many of which have been relegated to “dialect” status and excluded from orthographic development, schooling, and public life (Cutler & Dwyer, 1981; Leclerc, 2002; Singler, n.d.). These structural conditions predate displacement yet continue to shape refugee children’s access to education, emotional expression, family communication, and institutional support, rendering language both a marker of inequality and a critical pathway through which trauma exposure and recovery are ecologically mediated.

In displacement, many parents—already conditioned to view their heritage languages as inferior—found themselves unable to communicate fluently in English while also losing active competence in their native tongues. As children increasingly adopted Nigerian host languages, families experienced a form of subtractive multilingualism in which successive language acquisition eroded, rather than enriched, linguistic and emotional identity (Ethnolinguistic Identity Theory [ELIT]; Tajfel & Turner, 1979). Ecological disruptions in the refugee environment—including the breakdown of family support, limited access to schools, and minimal institutional or community support—further amplified these vulnerabilities (Ecological Systems Theory [EST]; Bronfenbrenner, 1979). These frameworks highlight that linguistic shifts and psychosocial outcomes are co-constructed: children’s language choices reflect identity negotiations while being constrained by systemic social and environmental factors.

Among refugee children, such conditions may result in language attrition and, in some cases, semi-lingualism, where neither the heritage language nor the dominant language of schooling is fully mastered (Karlander, & Salö,2026.; Skutnabb-Kangas, 1981; BAKER. 1993). Ethnolinguistic Identity Theory, derived from Social Identity Theory (Tajfel & Turner, 1979), helps explain how the devaluation of minority languages within unequal social hierarchies contributes to these patterns of language loss.

Within such environments, children may experience difficulties expressing emotions, narrating distress, or sustaining a stable connection to parental identity and cultural memory. From a social ecological perspective (Bronfenbrenner, 1979), these language-related disruptions may undermine family communication and identity continuity, thereby increasing vulnerability to psychosocial distress.

Among refugee children, disruptions in intergenerational language transmission frequently result in language attrition and forms of semilingualism, characterized by incomplete acquisition of both heritage languages and the dominant language of schooling. This linguistic precarity constrains affective articulation, narrative coherence, and the capacity to externalize distress, while weakening attachment to parental identity and cultural memory. Such disruptions fracture communicative reciprocity within families and amplify psychosocial vulnerability in displacement contexts.

This pattern is particularly salient in the Sierra Leonean case. Language historically fulfilled educational, communal, national, and nationist functions, with Krio operating as a near-universal lingua franca spoken by approximately 97% of the population (Fyle, 1994; Turay, 2022). However, the extreme violence of the civil war disrupted its integrative symbolic capacity, reindexing linguistic meaning from social cohesion to traumatic signification and leaving refugee children suspended between collective linguistic identity and the stigmatized semiotics of war.

Despite extensive research on war-related trauma among refugee populations, the mental health implications of linguistic disruption remain poorly understood among refugee children in Nigeria. Existing studies have primarily examined physical, sexual, and witnessed trauma as predictors of PTSD and psychological distress (Yarseah et al., 2025; Yarseah & Adegoroye, 2019), with limited attention to how language loss, semilingualism, and disrupted communication shape functional impairment or disturbances in self-organization associated with complex PTSD. Consequently, language remains largely absent from prevailing trauma models, despite its central role in emotional regulation, identity formation, and social functioning.

This study addresses this gap by examining how language experience—including heritage-language proficiency, home-language use, and host-language communication—interacts with trauma exposure to predict PTSD, complex PTSD, and functional impairment among stateless Liberian and Sierra Leonean refugee children in Nigeria. By integrating sociolinguistic and psychological perspectives, the study advances a socio-ecological understanding of trauma in contexts of protracted displacement.

### Problem Statement

Despite extensive evidence that armed conflict and displacement adversely affect children’s psychosocial well-being, few studies have examined the interplay of language, identity, and ecological systems among stateless Liberian and Sierra Leonean refugees in Nigeria. Many of these children are born into families without durable legal status due to failed resettlement, unfeasible repatriation, and restrictive naturalization policies, leaving them in prolonged legal limbo. This legal marginalization shapes children’s access to education, social services, and community supports, creating structural conditions that exacerbate trauma exposure and disrupt family and language environments. Prior research has documented intergenerational heritage-language erosion and shifts toward socially dominant languages among displaced populations (Fishman, 1966; Veltman, 1983). In refugee contexts, these processes are intensified by forced displacement, educational disruption, and trauma exposure, often resulting in reduced heritage-language proficiency, altered home-language practices, and uneven acquisition of host-country languages. Yet, the mechanisms through which language proficiency, language use, and emotional connection to language shape functional impairment associated with PTSD and CPTSD remain insufficiently examined.

Ethnolinguistic Identity Theory (Tajfel & Turner, 1979) suggests that heritage-language proficiency and use within the family may protect against identity disruption and relational breakdown, potentially reducing trauma-related functional impairment. Complementarily, Ecological Systems Theory (Bronfenbrenner, 1979) highlights how host-language competence facilitates engagement with educational, social, and institutional systems, while the collapse of family and community supports may exacerbate the psychological effects of trauma. Together, these perspectives suggest that the interaction between heritage-language maintenance, host-language use, and trauma exposure may be central to understanding variability in PTSD and CPTSD functional impairment among refugee children.

In the context of Oru Camp, both linguistic and ecological disruptions intersect: children face subtractive multilingualism, semi-lingualism, and weakened ties to parental and ethnic identity, while simultaneously navigating fractured microsystems, collapsed mesosystems, and diminished macrosystem support. These structural vulnerabilities mirror broader post-conflict and socio-economic challenges identified in Sierra Leone, including poverty, weak infrastructure, and limited livelihood opportunities (Horn et al., 2021), which align with ongoing gaps in SDG 4 (Quality Education), SDG 10 (Reduced Inequalities), and SDG 16 (Peace, Justice, and Strong Institutions). Such deficits exacerbate children’s exposure to trauma and constrain protective ecological supports in former refugee camp settings.

This dual vulnerability raises critical questions: To what extent does heritage-language competence moderate the psychological impact of trauma? How do systemic ecological factors interact with individual language practices to shape resilience and psychosocial outcomes? Addressing these gaps is essential for understanding how language and ecological contexts jointly influence identity, adaptation, and mental health in refugee populations and for informing culturally and linguistically sensitive interventions.

### The objectives of this study are to

1. Examine the association between native-language proficiency and PTSD- and CPTSD-related functional impairment among refugee children.
2. Assess whether patterns of native-language use in the home are associated with PTSD and CPTSD functional impairment.
3. Investigate the association between host-language communication, emotional connection to language, and functional impairment across PTSD and CPTSD outcomes.
4. Examine the interactive effects of heritage-language proficiency, parental/home-language use, and host-language (Nigerian) use on PTSD and CPTSD functional impairment.
5. Determine whether exposure to emotional trauma and sexual trauma predicts PTSD and CPTSD-related functional impairment.
6. Determine whether exposure to witnessed trauma and physical trauma predicts PTSD-and CPTSD-related functional impairment.

### Literature Review

Ethnolinguistic Identity Theory (ELIT), derived from Tajfel and Turner’s social identity theory, explains how language use shapes identity, particularly among socially subordinate groups (Tajfel & Turner, 1986). ELIT builds on four key concepts: social categorization (ingroup vs. outgroup distinctions), social identity (awareness of group membership), social comparison (favoring the ingroup over others), and psychological distinctiveness (a positive self-concept from group affiliation) (Giles &, Johnson, 2010). These processes influence how individuals define themselves in social contexts and extend specifically to the use and maintenance of language as a marker of ethnic identity.

Applied to Liberian and Sierra Leonean refugee children in Nigeria, ELIT clarifies why many children adopt English and Nigerian languages while heritage languages decline. English and Nigerian languages carry higher social prestige and utility, so children emphasize them to achieve social inclusion (social comparison), even as this weakens their ties to parental and ethnic identity (social identity). Parental attitudes further reinforce this process: when parents themselves struggle with or deprioritize their indigenous language, they implicitly signal to children that the language—and by extension, the associated ethnic identity—has limited value. Prior research in Oru Camp demonstrates that both parents and children increasingly relied on English and Nigerian Pidgin, linking language preferences closely to identity, belonging, and psychological adjustment (Nwagbo, 2016).

The study of language maintenance and erosion highlights that, although group-level tendencies are important, individuals actively make choices about which languages to use. ELIT provides a framework to understand these micro-level strategies, particularly in interethnic interactions. Some individuals accentuate ethnolinguistic characteristics (divergence) to maintain group identity, while others converge toward dominant languages to facilitate social integration (Giles &, Johnson, 2010). Divergence can be considered a form of short-term language maintenance, especially when societal norms favor out group languages and social sanctions exist for using the heritage language.

A central concept in ELIT is perceived vitality, referring to an individual’s perception of their ethnic group’s status, demographic strength, and institutional support for its language (Giles &, Johnson, 2010). Higher perceived vitality strengthens group identification and promotes the use of heritage language features. In the refugee context, perceived vitality is often low: local ethnic communities are small, connections to the homeland are disrupted, and institutional support is minimal. Coupled with parental reliance on socially dominant languages, reduced vitality diminishes ethnic belonging and encourages children to prioritize languages that are functional or socially valued.

This study introduces a novel theoretical perspective by positioning language as a moderating variable in the relationship between trauma exposure and psychosocial outcomes. To our knowledge, no prior research has examined this pathway in stateless refugee children. We argue that maintaining strong connections to a heritage language may reinforce cultural identity and act as a protective buffer against the psychological impact of trauma, whereas language attrition and semi-lingualism may weaken identity development, reduce adaptive capacity, and increase vulnerability to mental health challenges.

### Ecological Systems Theory (EST)

Bronfenbrenner’s Ecological Systems Theory (EST) depicts how interconnected levels of environment influence children’s development (Bronfenbrenner, 1977). Dissatisfied with developmental research that overlooked broader contexts, he proposed an ecological model as a common framework for understanding how nested systems shape outcomes (Shelton, 2019). Anderson et al (2004) further apply this lens to refugees, emphasizing how ecological contexts shift across stages of displacement.

The socioecological framework identifies four nested systems—microsystem, mesosystem, exosystem, and macrosystem—later expanded to include the chronosystem (Bronfenbrenner, 1977). This ecological lens situates trauma exposure and its consequences for PTSD, CPTSD and functional impairment within the layered realities of refugee life. Applied to the former Oru refugee camp in Ogun State, Nigeria, EST provides a framework for analyzing how the breakdown of social systems affected Liberian and Sierra Leonean refugees after the UNHCR cessation clause of 2012, which ended international protection and services.

At the microsystem level, families and peers provide primary support for refugee children, and interventions that strengthen family communication, particularly when culturally grounded and delivered by community members, improve child mental health outcomes (Betancourt et al., 2013). The mesosystem, which links families to schools and community resources, can be weakened when NGOs and UNHCR withdraw support, increasing social isolation (Peterson & Bush & 2012). At the exosystem level, restrictive government policies and limited integration programs indirectly constrain education, employment, and legal recognition, reinforced by host-community exclusion (Anderson et al., 2004). The macrosystem encompasses broader cultural and political forces, including Nigeria’s refugee policies and regional postwar fragility, while the **chronosystem** captures temporal shifts in support that produce intergenerational effects such as language loss, poverty, and reduced resilience (Anderson et al., 2004). Overall, risks in Oru camp are systemic, with the removal of exosystem and macrosystem supports amplifying vulnerabilities in fragile microsystems.

This study also acknowledges Bronfenbrenner’s later Process–Person–Context–Time (PPCT) model (Bronfenbrenner &, Morris,1998; 2006). PPCT highlights the importance of proximal processes and the role of time in shaping development. This is especially relevant for Oru, where nearly 90% of participants were born during or after the 2012 cessation. Here, the chronosystem is not just background context but a defining variable: it shapes an entire generation growing up without international protection, making time itself both a risk and resilience factor.

### PTSD and Complex PTSD Functional impairment

Extensive research indicates that exposure to trauma early in life has lasting negative effects on psychological well-being across the lifespan (Grasso, Ford, & Briggs-Gowan, 2012; Sharma, 2026; Fan, & Kang, 2025; van der Kolk, 2003). Children who experience traumatic events such as those commonly faced by migrant and refugee populations prior to migration and those living in refugee camp are at heightened risk for both immediate and long-term psychiatric symptoms (Shonkoff et al., 2012), including PTSD functional impairment such as deficits in attention, working memory, emotion regulation, and executive functioning, which can affect learning, social interactions, and daily activities (Fan & Kang, 2025; Dunn et al., 2017). Many of these pre-migration experiences meet Criterion A of the DSM-5 definition of PTSD, meaning they qualify as precipitating events for the development of the disorder (American Psychiatric Association, 2013).

Although post-traumatic stress disorder (PTSD) is a recognized consequence of trauma, far less is known about how trauma contributes to functional impairments in refugee children who are stateless and excluded from durable protection. Functional impairments capture the ways trauma undermines children’s capacity to attend school, regulate emotions, form relationships, and participate in daily life. PTSD often has an impact on children’s and adolescents’ social or educational spheres (Lewis et al, 2019), compromising the ability to perform and take part in important and desired daily aspects of life. Also in a to identify which specific DSM-5 post-traumatic stress symptoms (PTSS) have the greatest impact on functional impairment in trauma-exposed children and adolescents, Bartels et al., (2023) found that negative beliefs, emotional numbing, diminished interest, and concentration problems are the PTSD symptoms most strongly linked to functional impairment in trauma-exposed youth, with irritability also important in some populations.

Although youth adaptation following migration varies (Motti-Stefanidi & Masten, 2017), structural factors place refugee children at heightened risk. In refugee camps such as Oru, the absence of government services and institutional support, combined with exposure to violence and community distress, amplifies trauma and constrains access to education, healthcare, and social support. These conditions contribute to long-term functional impairment, affecting cognitive, emotional, and social development (Cohodes et al, 2021; Pollmann, Rakesh, & Fuhrmann, (2025 Shonkoff et al., 2021; Trent et al., 2019).

Although studies such as Thabet and Vostanis (2010) focus on children living in active war zones, their findings demonstrate a robust association between trauma exposure and depressive as well as post-traumatic stress disorder (PTSD) symptoms. These findings provide a useful conceptual framework for understanding the enduring psychological risks faced by children in former refugee camp settings, where exposure to trauma may persist through the residual effects of war, forced displacement, social marginalization, and intergenerational transmission of stress.

In such environments, children’s experiences of violence within homes and communities include both direct physical trauma and the witnessing of severe interpersonal violence, each posing significant threats to safety and bodily integrity. Evidence indicates that many youth exposed to family and community violence encounter such physically threatening events across multiple contexts and over prolonged periods (Graham-Bermann et al., 2009; Lupindo et al., 2025; Suie et al., 2021; Le et al., 2018). In former refugee camp settings, such as Oru, these exposures are further compounded by the legacies of armed conflict and displacement, placing children at heightened risk for psychological distress, PTSD symptoms, and disturbances in self-organization.

Estimates of trauma exposure among children and adolescents vary widely depending on context and methodology, but rates of witnessing community violence are consistently high, ranging from approximately 39% to 85%, while direct victimization has been reported in up to 66% of youth. Exposure to other forms of trauma, including sexual abuse and large-scale disruptive events, further compounds psychological risk physical abuse, motional neglect, sexual abuse, exposure to domestic violence and chronic stressors such as war and poverty (Dye, 2018; Cross et al., 2017). Also, Evidence suggests that witnessing trauma can result in post-traumatic stress symptoms, emotional dysregulation, and functional impairment comparable to outcomes associated with direct victimization (Ford et al., 2010).

In post-conflict and former refugee camp environments such as Oru, these cumulative exposures place children at heightened risk for persistent psychological distress, post-traumatic stress symptoms, and disturbances in self-organization, underscoring the need for trauma-informed research and intervention approaches.

Despite these risks, little research has systematically examined how different forms of trauma exposure shape multiple domains of functional impairment—such as emotional regulation, social functioning, and adaptive capacity—among stateless refugee children. This gap is critical, because without a clear understanding of the impairments that constrain adaptation and integration, interventions are likely to fall short in addressing the long-term needs of this highly vulnerable population.

Research has consistently shown that children exposed to neglect or raised in deprived or institutional environments are at heightened risk for delays in language development, with lasting consequences for academic achievement and emotional functioning (Naughton et al., 2013; Sala-Roca, 2019). Such delays reflect not only limited linguistic stimulation but also broader adverse conditions, including poverty, disrupted caregiving, and trauma exposure.

For Liberian and Sierra Leonean refugee children in Nigeria, these risks converge. Prolonged displacement, parental trauma, erosion of heritage language use, and limited access to quality education have created conditions in which children may neither fully acquire their mother tongue nor develop strong competence in English, resulting in language attrition and semilingualism. This linguistic vulnerability unfolds alongside chronic trauma exposure and structural deprivation, compounding risks for later difficulties in learning, emotional regulation, and social functioning.

### Language Development and Linguistic Vulnerability in Refugee Children

Building on this insight, the present study examines whether language use conditions the association between trauma exposure and mental health and functional outcomes among refugee children. Specifically, we investigate whether patterns of language dominance, attrition, and code-switching modify the strength of trauma-related impairment, thereby positioning language as a potential psychosocial buffer or risk factor in displaced children’s development.

Research on second-generation immigrant populations consistently demonstrates a strong shift toward the dominant societal language, with English proficiency and preference becoming nearly universal, while fluency in parental or heritage languages declines sharply across generations (Portes., & Hao, 1998). Importantly, heritage language retention is not uniform but varies substantially across groups, and these differences are not fully explained by individual or family-level factors alone. Also, Fishman (1966) and Veltman (1983) describe language shift as a three-generational process in which the first generation maintains the mother tongue at home, the second generation becomes bilingual with a growing preference for English in public domains, and the third generation loses effective knowledge of the parental language. In the context of Liberian and Sierra Leonean refugee children in Nigeria, this model is compressed: parents often have limited English proficiency due to Liberia’s English-only policy and Krio as a lingua franca in Sierra Leone, while children are exposed to English through schooling and Nigerian languages such as Yoruba or Igbo through socialization. These overlapping pressures—limited parental reinforcement of heritage language, English-medium education, and peer-language use—accelerate heritage language attrition and contribute to semi-lingualism among refugee children, highlighting the complex sociolinguistic dynamics that shape language acquisition in displaced populations.

Early research in the 20th century often portrayed bilingualism as a cognitive handicap, with studies such as Smith (1939) claiming that using two languages simultaneously caused speech delays in immigrant children, a view reinforced by the English-only ideologies of political figures like Theodore Roosevelt (Brumberg, 1986; Hakuta & Diaz, 1985). However, landmark studies by Peal and Lambert (1962), Leopold (1961), and Cummins (1978) demonstrated that fluent bilingual children exhibit greater cognitive flexibility, symbolic reasoning, and metalinguistic awareness. Large-scale sociological research further confirmed that bilingual students outperform limited bilinguals and monolingual peers academically, even after controlling for socio-economic status (Rumbaut, 1995; Portes and Rumbaut 1996). These findings indicate that bilingualism itself contributes to cognitive and academic development rather than merely reflecting innate ability (Hakuta & Diaz, 1985). For Liberian and Sierra Leonean refugee children in Nigeria, the challenge of semi-lingualism—driven by limited parental fluency, English-only schooling, and socialization in local Nigerian languages—is therefore structural and contextual, not cognitive, underscoring the importance of supporting heritage language maintenance alongside English acquisition.

While earlier scholarship framed linguistic outcomes among immigrant and refugee children primarily through cognitive or educational lenses, more recent interdisciplinary work situates language development within broader contexts of psychosocial adversity. For forcibly displaced children, language acquisition and use unfold alongside chronic exposure to violence, loss, and instability. Trauma exposure—whether through witnessing violence, direct physical or sexual victimization, or sustained emotional maltreatment—can disrupt neurocognitive, emotional, and social processes that are foundational to language development and communicative functioning. Accordingly, language must be understood not only as a developmental skill but also as a psychologically mediated process shaped by traumatic stress.

In Oru, stateless Liberian and Sierra Leonean refugee children experienced not only structural deprivation but also linguistic exclusion, as the absence of mother-tongue education limited both their academic development and cultural identity formation. According to UNESCO (2003), language is central to realizing the right to education and ensuring meaningful participation in society. This implies that linguistic marginalization may intensify trauma-related functional impairments among refugee children, who already face psychological and social vulnerabilities stemming from displacement and exposure to violence.

Exposure to trauma in childhood can have profound effects on neurocognitive and language development, particularly among refugee populations. Chronic or severe trauma disrupts attention, memory, executive functioning, and other cognitive systems that support communication, learning, and psychosocial adaptation (Malarbi et al., 2017; Shonkoff et al., 2012). These disruptions may undermine higher-order language skills, including narrative organization, autobiographical memory, and social communication, which are critical for everyday functioning and academic engagement (Gillam & Hyter, 2025). Consequently, children with trauma histories may experience functional impairment not only through emotional dysregulation but also through challenges in language-mediated processes. When stress is chronic and/or severe, the body’s stress response system may remain activated, a condition known as toxic stress (Nelson et al., 2020). Toxic stress can disrupt regulatory systems, leading to prolonged physiological arousal that may alter, or even permanently damage, the brain. These persistent changes can result in “permanent changes to the body, brain, and mind” (Hyter et al., 2024) and often impair developmental processes, including language and communication (Hyter et al, 2024; Murphy et al., 2022).

Studies further suggest that exposure to physical, witnessing, and emotional trauma disrupts not only basic language acquisition but, more critically, higher-order communicative functions that rely on emotional regulation, social cognition, and memory. Children exposed to trauma—including those who primarily witness violence or experience chronic emotional stress—consistently demonstrate difficulties with conversational turn-taking, understanding language in emotionally laden contexts, and expressing ideas coherently (Hyter et al, 2024). Reviews of the literature further indicate that while basic language skills may be affected, the most pronounced impairments occur at the level of conversational and discourse language, which are essential for social interaction and everyday functioning (Sylvestre et al., 2016). These language disruptions appear to be mediated by trauma-related impairments in executive functioning, working memory, and perspective-taking abilities, particularly in children exposed to prolonged or repeated trauma (Hyter, 2021). Importantly, evidence from discourse-focused studies shows that children with maltreatment histories may perform adequately on surface-level language tasks yet struggle with narrative and expository discourse, with older children showing greater impairment—suggesting cumulative effects of chronic trauma exposure over time (Ciolino et al., 2021). This pattern is especially relevant for refugee children whose trauma exposure often involves witnessing violence, persistent emotional insecurity, and physical threat rather than isolated incidents of direct victimization.

Emotion is conveyed through all aspects of language (Majid, <u>2012</u>), but there are some language that has more emotion than others (Fatima et al., 2024). According to Fatima et al., (2024), Languages often become emotionally associated with some particular life experiences, relationships, and cultural settings like the experience of living in disbanded refugee camp without international protection. In this instant, speakers may feel more comfortable expressing vulnerability in their first language, associating it with danger and abandonment, while second or third languages may be used in more neutral or formal emotional contexts (Safiullin & Zheltukhina, 2020), a phenomenon is known as “emotional resonance,” where the emotional impact of a language is shaped by cultural associations and personal history (Costa, 2021). Language has power and this power influences people emotion making them to swift from one language to another. For multilingual, switching between languages can sometimes imply switching emotional connections, highlighting the malleability and fluidity of emotional experiences across linguistic boundaries (Serrano et al., 2020). Research has indicated that language affects not only the intensity of emotions but also the ease or difficulty of expressing them (Llanes, 2020). Also, recent research highlights the intricate relationship between language and emotion, demonstrating that language not only conveys emotional content but also shapes emotional experience, perception, and regulation (Ferré, Fraga, & Hinojosa, 2025).

Various studies state that language influences emotional development from an early age (Ponari et al., 2020; Sabater et al., 2023) and that infants with stronger language skills are better at identifying emotions (Nook et al., 2017; Ruba et al., 2021). In the same vain, speaking a non-native language can lead to a feeling of emotional detachment, often referred to as the “foreign language effect” (Aguilar, Ferré., & Hinojosa, 2024; Ferré., Fraga, & Hinojosa, 2025; Caldwell-Harris, 2014).

Another area of research has focused on examining the influence of language on emotion processing. Although no empirical studies have directly examined the relationship between emotional connection to language on PTSD or CPTSD-related functional impairment among refugee children, existing scholarship points to plausible pathways through which language practices may shape psychological functioning. This perspective provides evidence for the role of language in acquiring, experiencing, understanding and regulating emotions. Language gives children an additional means of communicating their needs and of understanding their own and others’ emotions. For example, infants with higher language abilities are better at regulating their emotions when frustrated (Fields-Olivieri et al., 2024; Pemberton-Roben et al., 2012). Additionally, affect labelling, a form of implicit emotion regulation, has been shown to increase positive feelings when naming positive events (Vlasenko et al., 2021) and reduce negative feelings when naming negative events (Niles et al., 2015).

More so, factor that might determine choice of language is positive emotional feelings associated with the situation to be expressed. Multilinguals tend to select languages according to the emotional and social context in which they operate. In emotionally intimate situations—such as expressions of love, grief, or anger—individuals are more likely to use a language with strong personal significance, typically their first language (Schwieter & Sunderman, 2020), as it often carries the deepest emotional resonance (Fatima et al., 2024). Conversely, in public or socially evaluative contexts, multilingual speakers may choose a language perceived as more neutral, socially powerful, or widely accepted. Among Liberian and Sierra Leonean refugee children in the Oru community, Yoruba or Igbo may serve this instrumental function due to their broader societal utility. However, when heritage language use declines—particularly in families where parents themselves have experienced language attrition—children may have fewer opportunities to develop strong emotional attachment to their heritage language, potentially affecting emotional expression and identity formation.

Research among heritage bilinguals indicates that heritage-language (HL) proficiency plays a significant role in acculturation outcomes, with family language practices closely associated with cultural involvement, self-esteem, and psychological wellbeing (Lam & Catto, 2023). A substantial body of literature has further demonstrated strong links between HL use and heritage identity among ethnolinguistic minorities (Arrendondo et al., 2016; Kang, 2013; Shin, 2016).

However, less is known about how HL proficiency and everyday language practices relate to broader processes of acculturation and psychological adjustment, particularly in contexts of forced migration (Lindner et al., 2020; Yağmur & van de Vijver, 2012). While recent evidence suggests that HL maintenance uniquely contributes to self-esteem and wellbeing, little research has examined whether these linguistic and acculturative factors also predict trauma-related functional impairment among refugee children exposed to displacement and conflict-related stressors.

A heritage language (HL) typically refers to a language other than that of the host country tied to a speaker’s family or ancestral homeland, varying in personal relevance (Scontras et al., 2015; Shin, 2016). HL use supports ethnic identity formation through family interactions and community engagement (Kang, 2013). Effective parent–child communication in the HL fosters secure attachments and positive outcomes such as self-esteem (Müller et al., 2019). Conversely, HL attrition can lead to family misunderstandings, community alienation, identity confusion, and reduced self-esteem (Brown, 2009; Chen & Padilla, 2019; Liu et al., 2009). More importantly, research on Syrian refugee children in Türkiye shows that second-language barriers not only hinder academic performance but intersect with trauma, belonging, and peer exclusion, thereby compounding psychosocial vulnerability (Aksaç, 2025). These patterns suggest that heritage-language maintenance may protect psychosocial adjustment, whereas attrition may exacerbate vulnerabilities, which can be particularly consequential for trauma-exposed refugee children.

Research on refugee language outcomes suggests that age at displacement plays a critical role in language development. Studies of refugee populations in Western host countries consistently show that individuals who arrive at younger ages demonstrate better language proficiency outcomes than those displaced later in life, highlighting the existence of sensitive developmental periods for language acquisition (Van Tubergen,2010). Also, Research on immigrant integration consistently shows that host-country language proficiency shapes labor market outcomes, social integration, and the educational trajectories of children (Chiswick & Miller, 2002; Martinovic et al., 2009; Shields & Price, 2002). Beyond economic consequences, these studies suggest that language proficiency functions as a critical resource for intergenerational adjustment.

Fridrikh, Pentón Herrera, and Kałdonek-Crnjaković (2025) found that multilingual strategies, particularly proficiency in a commonly shared or emotionally accessible language, support social integration and emotional well-being among Ukrainian refugee children in Polish schools. In the context of Liberian and Sierra Leonean refugee children in Nigeria, higher proficiency in both heritage and host languages may facilitate social interaction, emotional expression, and support-seeking, thereby reducing functional impairment associated with PTSD and CPTSD . Conversely, limited proficiency—especially when parents themselves have experienced heritage-language attrition—may exacerbate emotional detachment and functional difficulties, aligning with the “foreign language effect”. These considerations underscore the relevance of language proficiency not only for communication but also for emotional regulation and psychosocial adaptation. Also, UNESCO (2025) emphasizes that supporting multilingualism in refugee education—including access to mother-tongue instruction alongside second-language learning—can reduce trauma and foster a stronger sense of belonging. Similarly, Becker and Magno (2022) highlight that inclusive language policies not only facilitate linguistic integration but also promote psychosocial well-being among displaced students. These findings suggest that refugee children’s proficiency in both heritage and host languages may buffer functional impairment associated with PTSD and CPTSD by supporting emotional expression, social interaction, and access to social support.

Language proficiency plays a critical role in shaping emotional expression among multilingual individuals. Higher proficiency in a given language facilitates the articulation of complex emotions due to a richer emotional vocabulary and deeper familiarity with cultural norms (Safiullin & Zheltukhina, 2020; Caldwell-Harris et al., 2022). Conversely, limited proficiency can constrain emotional expression, particularly in non-native languages. For refugee children, this effect is compounded when parents themselves have limited heritage-language proficiency and primarily communicate in English or Liberian Pidgin (Singler, 2006; DeCamp, 2012). These dynamics suggest that both parental and child language proficiency may influence children’s ability to express and process emotions, potentially affecting PTSD- and CPTSD-related functional impairment.

### Ethical Considerations

Ethical approval for the study was obtained on 26 February 2026 from the Ethical Committee for Research, Development, and Innovation at Ekiti State University (Reference ORDI/EKSU/EAC/26/025). Data collection was conducted between 27 February 2026 and 10 March 2026. Written informed consent was obtained from parents or legal guardians, and verbal or written assent was obtained from all participating children. Participants and their guardians were informed, in age-appropriate language, about the study purpose, procedures, confidentiality, voluntary participation, and the right to withdraw at any time without penalty. All data were anonymized using participant ID codes, securely stored in locked cabinets and password-protected files, and accessible only to authorized research team members. Questionnaires were administered in private settings to ensure confidentiality. All procedures adhered to the Helsinki Declaration and principles of respect, beneficence, and protection of vulnerable populations.

## Materials and Methods

### Study Design

This was a cross-sectional study examining trauma exposure, language use, and PTSD/CPTSD functional impairment among refugee children and adolescents residing in the Oru refugee camp in Nigeria. The cross-sectional design enabled the assessment of relationships between trauma experiences, social support, and language use at a single time point, providing an overview of risk and protective factors affecting mental health and functional outcomes.

### Census of the Accessible Population

The refugee settlement hosts approximately 400–500 children. For this study, the population was restricted to children aged 6–17 years who were enrolled in settlement schools and met the inclusion criteria, resulting in an accessible population of 320 eligible participants. The minimum required sample size was calculated using Cochran’s formula at a 95% confidence level, a 5% margin of error, and an assumed population proportion of 0.5, yielding an initial estimate of 384 participants for an infinite population. After applying the finite population correction for N = 320, the required sample size was approximately 175 participants. Given that the accessible population was relatively small and manageable, a census approach was adopted, inviting all 320 eligible participants to maximize coverage and representativeness of the accessible population.

### Measures

#### Child and Adolescent Trauma Screen (CATS)

The Child and Adolescent Trauma Screen (CATS; Sachser et al., 2017) was developed to screen for exposure to potentially traumatic events (PTEs) and posttraumatic stress disorder (PTSD) symptoms in children and adolescents. The CATS is a self-report instrument assessing potentially traumatic events (15 items), DSM-5 PTSD symptoms (20 items), and psychosocial functioning (5 items) for children aged 7–17. Items are rated on a four-point scale (0 = never to 3 = almost always), with total PTSD scores ranging from 0 to 60. A cut-off score of ≥21 was used to indicate clinically significant symptoms. An international validation study demonstrated good to excellent reliability for the measure, with α ranging from .88 to .94 (Sachser et al., 2017). Convergent-discriminant validity was supported, with medium to strong correlations with depression (r = .62–.82) and anxiety (r = .40–.77), and low to medium correlations with externalizing symptoms (r = −.15–.43) across informants in all language versions. Confirmatory factor analysis (CFA) supported the underlying DSM-5 four-factor structure—reexperiencing, avoidance, negative alterations in mood and cognition, and hyperarousal—using samples of n = 475 for self-report and n = 424 for caregiver reports.

#### International Trauma Questionnaire – Child/Adolescent (ITQ-CA)

The ITQ-CA is a 22-item self-report instrument assessing ICD-11 PTSD and Complex PTSD (CPTSD) in children and adolescents aged 7–17 years (Cloitre et al., 2018). It consists of six PTSD items across three symptom clusters (reexperiencing, avoidance, and sense of current threat) and six CPTSD items assessing disturbances in self-organization (affective dysregulation, negative self-concept, and interpersonal difficulties), with additional items assessing functional impairment across social, academic, and family domains.

The International Trauma Questionnaire Child/Adolescent Version (ITQ-CA) has demonstrated strong discriminant validity across multiple cultural contexts. Validation studies indicate that the measure reliably differentiates between young people meeting criteria for PTSD alone and those meeting criteria for CPTSD (Haselgruber et al., 2020a; Ho et al., 2022; Kazlauskas et al., 2020; Redican et al., 2022). Specifically, individuals with PTSD alone exceed the diagnostic threshold on PTSD symptom clusters without meeting criteria for disturbances in self-organization (DSO), whereas those with CPTSD exceed thresholds on both PTSD and DSO subscales. The ITQ-CA further incorporates functional impairment items related to social, academic, and family domains, ensuring comprehensive assessment consistent with ICD-11 diagnostic requirements.

Responses are rated on a five-point Likert scale (0 = “never” to 4 = “almost always”), allowing both categorical diagnostic scoring and dimensional measurement of symptom severity. In the current study, the ITQ-CA functional impairment subscale demonstrated good reliability (α = 0.83, CR = 0.86, AVE = 0.54). The AVE exceeded the recommended threshold of 0.50, supporting convergent validity, and HTMT ratios were below 0.85, confirming discriminant validity. These findings indicate that the functional impairment construct was measured reliably within this sample (Fornell, & Larcker, 1981).

##### Sociolinguistic Questionnaire

A brief questionnaire developed for the study assessed language use and proficiency, including language spoken at home, use of Nigerian languages, native dialect exposure, comprehension, and self-rated speaking ability. Internal consistency for the composite language items was acceptable (Cronbach’s α = 0.74; CR = 0.78; AVE = 0.52), supporting reliability and convergent validity. HTMT ratios with PTSD and CPTSD measures were below 0.85, indicating discriminant validity. These variables were treated as categorical predictors in analyses examining functional impairment across PTSD and CPTSD groups.

#### Method of Data Analysis

The dataset was screened for completeness, missing values, and multivariate outliers using Mahalanobis distance (p < 0.001), with extreme cases removed. MANOVA assumptions were checked: homogeneity of variance-covariance matrices with Box’s M test and equality of variances for individual dependent variables using Levene’s test, both confirming acceptable conditions. Descriptive statistics (means, standard deviations, frequencies, percentages) summarized demographics, trauma exposure, language proficiency, and functional impairment. Cross-tabulations with chi-square tests examined relationships between categorical variables such as gender, age, and language proficiency. MANOVA was conducted to assess group differences in PTSD and CPTSD functional impairment across trauma exposure and language proficiency levels. Multivariate effects were reported using Roy’s Largest Root, Pillai’s Trace, Wilks’ Lambda, and Hotelling’s Trace, with effect sizes interpreted using partial eta squared (η²). Post-hoc analyses identified specific group differences where multivariate effects were significant. All analyses were performed in IBM SPSS Statistics, version 24 with a significance threshold of α = 0.05.

## 3.0 Demographic Characteristics

### 3.0 Demographic Characteristics

The study included 320 refugee children aged 6–17 years residing in the Oru settlement. Participants were slightly more female (57.2%) than male (42.8%) and were relatively evenly distributed across age groups: 6–10 years (34.4%), 11–13 years (31.3%), and 14–17 years (34.4%), with a mean age of 12.0 years (SD = 2.91). Most children were of Liberian origin (56.3%), with the remainder from Sierra Leone (43.8%). Educational enrollment spanned primary (42.2%), junior secondary (32.8%), and senior secondary levels (25.0%), reflecting broad school participation across developmental stages. Overall, the demographic profile represents a diverse cohort of school-aged refugee children who have spent substantial portions of their formative years in displacement.

The sample comprised 320 stateless refugee children with a moderately balanced gender distribution (42.8% males, 57.2% females). Participants ranged in age from 6 to 17 years (M = 12.0, SD = 2.91), with a relatively even distribution across age groups. Children aged 6–10 years and 14–17 years each constituted 34.4% of the sample, while those aged 11–13 years represented 31.3%.

In terms of country of origin, a slight majority of participants were from Liberia (56.3%), with the remainder originating from Sierra Leone (43.8%), reflecting the historical composition of the displaced population in the camp. Educational enrollment was distributed across primary (42.2%), junior secondary (32.8%), and senior secondary levels (25.0%), indicating broad school participation across developmental stages. Overall, the demographic profile reflects a diverse cohort of school-aged refugee children who have spent substantial portions of their formative years in displacement.

Table 2 summarizes patterns of language exposure, proficiency, emotional connection, and language use among refugee children (N = 320). Overall, the distribution indicates a clear shift away from heritage (L1) languages toward English and Nigerian languages. Heritage-language exposure at home was inconsistent: one-third of participants (33%) reported that parents or grandparents never spoke their dialect at home, while only 39% reported that this occurred often or always. Similarly, 64% reported that parents never or only sometimes used their native language when speaking to each other, suggesting limited intergenerational transmission of L1.

**Table 1.** Demographic Characteristics of Refugee Children (N = 320)

| Variable | Category | n | % |
| --- | --- | --- | --- |
| **Gender** | Male | 137 | 42.8 |
|  | Female | 183 | 57.2 |
| **Age Group** | 6–10 years | 110 | 34.4 |
|  | 11–13 years | 100 | 31.3 |
|  | 14–17 years | 110 | 34.4 |
| **Mean Age (SD)** | — | 12.0 | 2.91 |
| **Country of Origin** | Liberia | 180 | 56.3 |
|  | Sierra Leone | 140 | 43.8 |
| **Education Level** | Primary | 135 | 42.2 |
|  | Junior Secondary | 105 | 32.8 |
|  | Senior Secondary | 80 | 25.0 |

**Table 2:**
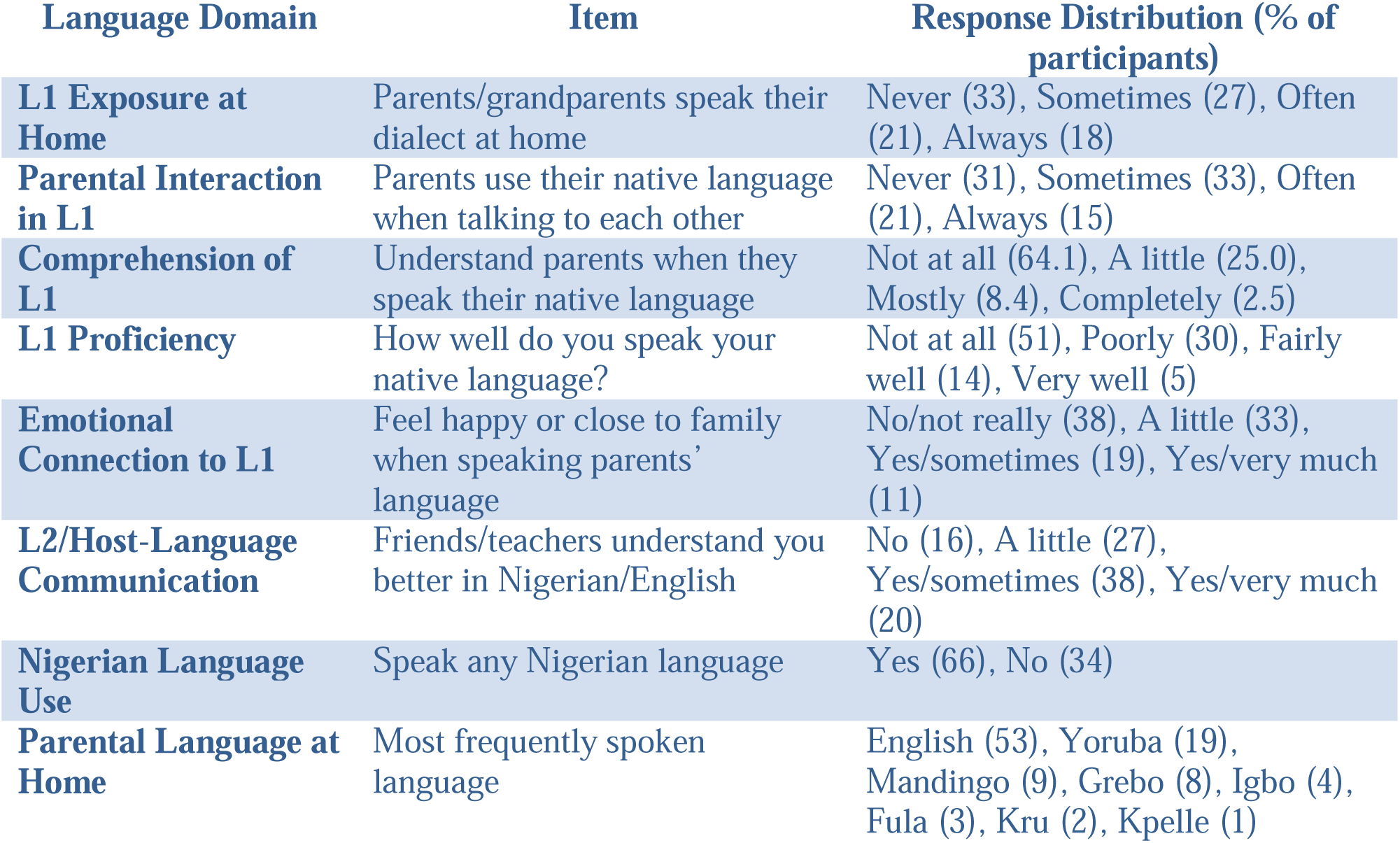
Language use and proficiency among Refugee Children (N=320)

Comprehension and productive proficiency in L1 were generally low. Nearly two-thirds of participants (64.1%) reported no understanding when parents spoke their native language, and only 10.9% reported mostly or complete comprehension. Expressive proficiency was even more limited, with 81% reporting that they spoke their native language not at all or poorly, and only 5% indicating very good proficiency. These distributions indicate substantial erosion of functional heritage-language competence. Despite low proficiency, emotional connections to L1 were partially retained. While 38% reported no emotional connection to their parents’ language, 63% endorsed at least some degree of emotional closeness when using it, suggesting that affective ties to heritage languages may persist even in the absence of strong linguistic ability.

In contrast, host-language communication was comparatively strong. Over half of participants (58%) reported being understood sometimes or very much by friends or teachers in Nigerian or English, and two-thirds (66%) reported speaking at least one Nigerian language. English emerged as the dominant home language (53%), with heritage languages such as Yoruba, Mandingo, Grebo, and Igbo spoken by smaller proportions. Collectively, these findings depict a pattern of language shift characterized by limited L1 exposure and proficiency alongside greater functional reliance on English and Nigerian languages.

Exploratory descriptive comparisons by religious affiliation suggested differences in home-language patterns. Among Christian households, English was most frequently reported as the primary home language (approximately 53%). In contrast, Muslim households reported greater use of Yoruba (26%) and Fula (14%), while children from African Traditional Religion backgrounds showed a more balanced distribution across Fula (15%), Krun (15%), and Grebo (12.5%). Overall, these patterns indicate a broader trend of linguistic assimilation alongside socially patterned variation in heritage-language use across religious groups.

As shown in Table 3, there were notable differences in the primary language spoken at home across religious groups. Among Christian households, English was the dominant home language (52.8%), followed by Yoruba (17.2%) and Grebo (8.9%). In contrast, among Muslim families, English was less common (32%), with a higher proportion speaking Yoruba (26%) and Fula (14%). Refugee children from African Traditional Religion backgrounds showed the lowest use of English (10%), with more balanced use of Fula (15%), Krun (15%), and Grebo (12.5%).

**Table 3:** Primary Language Spoken at Home by Religion (N = 320)

| Language Spoken at Home | Christianity (n=180) | Islam (n=100) | African Traditional Religion (n=40) | Total (%) |
| --- | --- | --- | --- | --- |
| **English** | 95 (52.8%) | 32 (32.0%) | 4 (10.0%) | 131 (40.9%) |
| **Yoruba** | 31 (17.2%) | 26 (26.0%) | 4 (10.0%) | 61 (19.1%) |
| **Grebo** | 16 (8.9%) | 9 (9.0%) | 5 (12.5%) | 30 (9.4%) |
| **Fula** | 8 (4.4%) | 14 (14.0%) | 6 (15.0%) | 28 (8.8%) |
| **Krun** | 6 (3.3%) | 8 (8.0%) | 6 (15.0%) | 20 (6.3%) |
| **Mandingo** | 13 (7.2%) | 8 (8.0%) | 3 (7.5%) | 24 (7.5%) |
| **Pidgin/Other Nigerian** | 11 (6.1%) | 3 (3.0%) | 2 (5.0%) | 16 (5.0%) |
| **Total** | **180 (100%)** | **100 (100%)** | **40 (100%)** | **320 (100%)** |

**Note.** Percentages are calculated within religious groups. “Primary language spoken at home” refers to the language most frequently used in daily family interactions. “Pidgin/Other Nigerian” includes Nigerian Pidgin and minority Nigerian languages reported by fewer than 5% of respondents.

Table 4 presents parental home language by age group, revealing a progressive shift toward English with increasing age. English use rises from 33% among 6–8-year-olds to 55% among 15–17-year-olds. In contrast, heritage languages such as Grebo, Krun, and Mandingo remain low or decline across age groups, while Yoruba use gradually increases from 17% to 23%, indicating partial adoption of the host language. These patterns reflect age-related heritage-language attrition and suggest that older children are at greater risk of semi-lingualism, exhibiting limited proficiency in both heritage and host languages. Overall, the findings highlight how developmental stage interacts with language exposure to shape bilingual competence and indicate potential risks for heritage-language loss in contexts of protracted displacement.

**Table 4:** Parental Home Language by Age Group (N = 320)

| Language | 6–8 yrs | 9–11 yrs | 12–14 yrs | 15–17 yrs | Total (%) |
| --- | --- | --- | --- | --- | --- |
| **English** | 18 (33%) | 25 (31%) | 40 (46%) | 48 (55%) | 131 (41%) |
| **Yoruba** | 9 (17%) | 14 (17%) | 18 (21%) | 20 (23%) | 61 (19%) |
| **Grebo** | 8 (15%) | 7 (9%) | 8 (9%) | 7 (8%) | 30 (9%) |
| **Fula** | 5 (9%) | 6 (8%) | 8 (9%) | 9 (10%) | 28 (9%) |
| **Krun** | 4 (7%) | 4 (5%) | 6 (7%) | 6 (7%) | 20 (6%) |
| **Mandingo** | 5 (9%) | 6 (8%) | 7 (8%) | 6 (7%) | 24 (8%) |
| **Other** | 5 (9%) | 9 (11%) | 8 (9%) | 6 (7%) | 28 (8%) |
| **Total** | **54** | **81** | **95** | **102** | **320** |

**Table 5:**
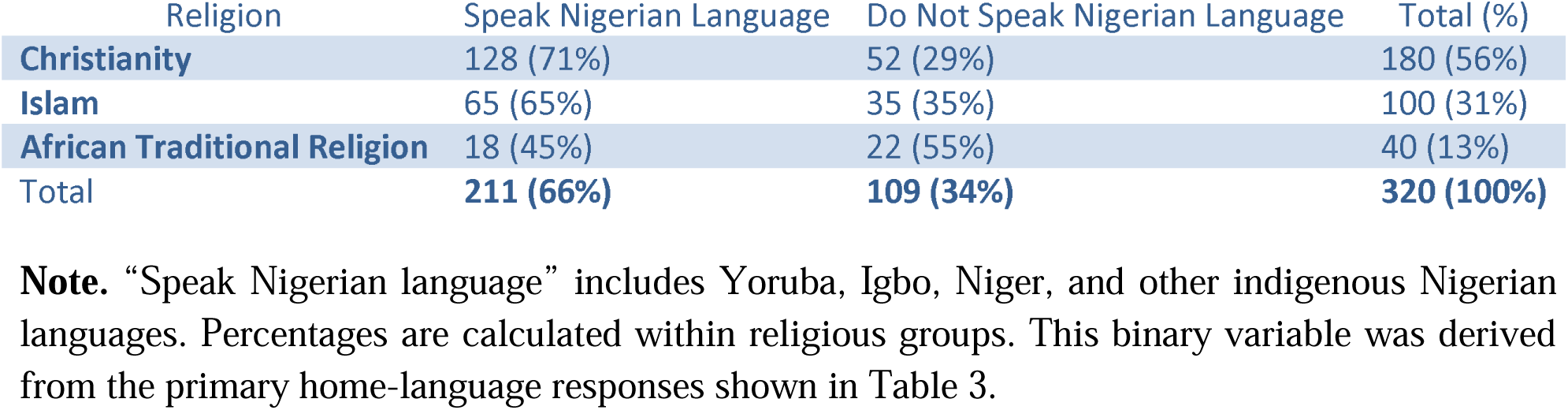
Binary Classification of Nigerian Language Use by Religion among Refugee Children (N = 320)

There was a significant association between religion and speaking a Nigerian language, χ²(2, N = 320) = 18.82, p = .001. Children from Christian (71%) and Muslim (65%) households were more likely to speak a Nigerian language than those from African Traditional Religion households (45%). These results indicate that the likelihood of speaking a Nigerian language varies across religious groups.

### 3.4 Relationship between Age and Nigerian Language Speaking

A chi-square test of independence indicated a statistically significant association between age group and speaking a Nigerian language, χ²(10, N = 320) = 19.37, p = .036. This result suggests that the likelihood of speaking a Nigerian language varies across age groups. Assumption checks indicated that the expected cell counts were adequate for chi-square analysis.

A chi-square test of independence indicated a statistically significant association between age group and speaking a Nigerian language, χ²(4, N = 318) = 13.27, p = .010, Cramér’s V = .20, suggesting a small-to-moderate association. Examination of row percentages (Table 6) showed that the proportion of children who spoke a Nigerian language was highest in younger age groups (6–8 years, 100%) and declined among older adolescents (15–17 years, 73.6%), with intermediate values in the middle age groups. A linear-by-linear association test confirmed a significant negative trend, χ²(1) = 3.98, p = .046, indicating that older children were less likely to speak Nigerian languages. One cell contained a zero count, so results should be interpreted with caution. Directional measures supported this pattern (Somers’ d = −.143, p = .043), reflecting a weak but significant negative association between age and Nigerian-language use. Other measures of association were very small, consistent with the modest strength of the relationship.

**Table 6:** Nigerian Language Speaking by Age Group (N = 318)

| Age Group<br>(years) | Speak Nigerian<br>Language (Yes) | Do Not Speak<br>(No) | Row %<br>Yes | Row %<br>No | Total<br>(n) | % of<br>Total |
| --- | --- | --- | --- | --- | --- | --- |
| 6–8 | 28 | 0 | 100.0% | 0.0% | 28 | 8.8% |
| 9–10 | 29 | 12 | 70.7% | 29.3% | 41 | 12.9% |
| 11–12 | 45 | 7 | 86.5% | 13.5% | 52 | 16.4% |
| 13–14 | 60 | 16 | 78.9% | 21.1% | 76 | 23.9% |
| 15–17 | 89 | 32 | 73.6% | 26.4% | 121 | 38.1% |
| Total | 251 | 67 | — | — | 318 | 100% |

**Notes:**
- Row % = proportion of each age group speaking / not speaking Nigerian language. - % of Total = proportion of total N in each age group. - N = 318 after excluding miscoded responses (“2”). - This table is fully consistent with $\chi^2(4, N = 318) = 13.27, p = .010$ .

Table 7 shows children’s self-reported understanding of their parents’ native language across age groups. Overall, most children reported being able to understand their parents’ language either often (32%) or always (28%), while smaller proportions indicated understanding sometimes (27%) or never (13%). Percentages within each age group were generally similar, indicating that comprehension did not vary substantially with age.

**Table 7:** Understanding Parents’ Native Language by Age Group (N = 320)

| Understanding Level | 6–8 yrs<br>(n=60) | 9–11 yrs<br>(n=81) | 12–14 yrs<br>(n=95) | 15–17 yrs<br>(n=84) | Total<br>(n=320) | Total<br>(%) |
| --- | --- | --- | --- | --- | --- | --- |
| **Never** | 7 (11.7%) | 10 (12.3%) | 11 (11.6%) | 14 (16.7%) | 42 | 13.1% |
| **Sometimes** | 15<br>(25.0%) | 21 (25.9%) | 24 (25.3%) | 26 (30.9%) | 86 | 26.9% |
| **Often** | 18<br>(30.0%) | 28 (34.6%) | 32 (33.7%) | 24 (28.6%) | 102 | 31.9% |
| **Always** | 20<br>(33.3%) | 22 (27.2%) | 28 (29.5%) | 20 (23.8%) | 90 | 28.1% |
| **Total** | 60 (100%) | 81 (100%) | 95 (100%) | 84 (100%) | 320 | 100% |

**Notes:**
1. Row % = proportion of each age group in that category. 2. Total (%) = proportion of total sample in each understanding level. 3. N = 320 after excluding any miscoded responses. 4. Counts are adjusted so the sum matches total N = 320.

A chi-square test of independence confirmed this pattern: there was no significant association between age group and understanding of parents’ native language, χ²(15, N = 320) = 9.72, p = .837. This result indicates that children across all age groups demonstrated comparable levels of understanding, suggesting that exposure and comprehension of parental native language remain relatively stable throughout childhood and adolescence.

### 3.8 Language Spoken at Home by Gender

χ²(7, N = 320) = 25.12, *p* = .001.

Table 8 shows the distribution of home languages among male and female refugee children. English was the most frequently reported home language for both males (45%) and females (38%), followed by Yoruba (19% for both genders). Other languages demonstrated gender-based differences: males were more often reported in Grebo-speaking households (12% vs. 7%), whereas females were more often reported in Fula (10% vs. 7%), Krun (8% vs. 4%), and Mandingo (9% vs. 6%) households. Counts and percentages for other less common languages, such as “Other,” were similar between genders.

**Table 8:**
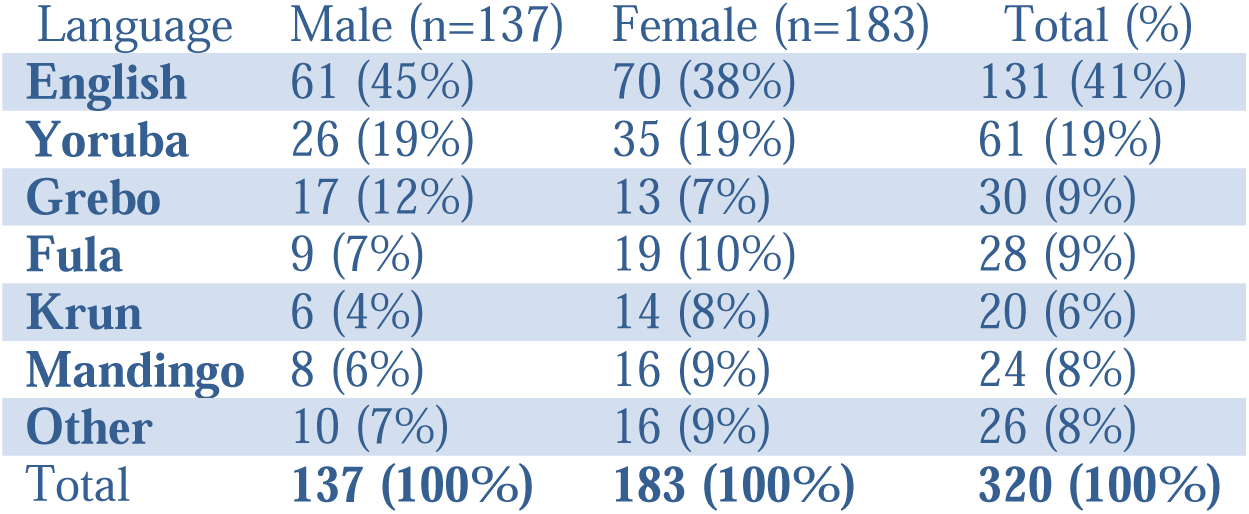
Home Language by Gender (N = 320)

A chi-square test of independence indicated a statistically significant association between gender and home language, χ²(7, N = 320) = 25.12, p = .001, suggesting that the linguistic composition of households differs by gender. Overall, these results indicate gender-based variations in the home-language environment among refugee children.

### Rationale for Focusing on Functional Impairment

The study focused on PTSD and CPTSD functional impairment rather than total symptom counts, in line with ICD-11’s emphasis on impairment as a core diagnostic criterion. Functional impairment provides a more ecologically valid indicator of trauma impact, capturing disruptions in schooling, emotional regulation, and interpersonal relationships among refugee children.

### Inferential Statistics

#### Hypothesis 1

Native-language proficiency would be associated with functional impairment among refugee children, such that higher proficiency would correspond to lower PTSD and CPTSD functional impairment.To test Hypothesis 1, descriptive statistics, multivariate analyses, and parameter estimates were examined (Tables 9)

#### Descriptive statistics

**Table 9** presents the mean and standard deviation of PTSD and CPTSD functional impairment across levels of native-language proficiency. PTSD-related functional impairment was lowest among children who reported speaking their native language *very well* (M = 2.00, SD = 1.84) and highest among those who reported speaking it *poorly* (M = 3.23, SD = 1.27). Children who did not speak their native language at all (M = 3.01, SD = 1.49) and those who spoke it fairly well (M = 2.93, SD = 1.54) reported intermediate levels of PTSD-related impairment.

**Table 9:** Descriptive Statistics for PTSD and CPTSD Functional Impairment by Native Language Proficiency (N = 320)

| Native Language Proficiency | CPTSD Functional Impairment (M ± SD) | PTSD Functional Impairment (M ± SD) |
| --- | --- | --- |
| Not at all | 3.14 ± 2.13 | 3.01 ± 1.49 |
| Poorly | 3.84 ± 1.75 | 3.23 ± 1.27 |
| Fairly well | 4.24 ± 1.32 | 2.93 ± 1.54 |
| Very well | 3.65 ± 2.15 | 2.00 ± 1.84 |
| Total | 3.53 ± 1.97 | 3.01 ± 1.47 |

CPTSD-related functional impairment showed greater variability across proficiency levels. The highest mean CPTSD impairment was observed among children who spoke their native language fairly well (M = 4.24, SD = 1.32), followed by those who spoke it poorly (M = 3.84, SD = 1.75). Lower mean levels were observed among children who spoke their native language very well (M = 3.65, SD = 2.15) or not at all (M = 3.14, SD = 2.13).

#### Multivariate analysis

A multivariate analysis of variance (MANOVA) was conducted with PTSD and CPTSD functional impairment as dependent variables and native-language proficiency as the independent variable, controlling for country of origin. Assumptions for MANOVA were met.

The multivariate effect of native-language proficiency was statistically significant, Pillai’s Trace = .082, F(6, 630) = 4.48, p < .001, partial η² = .041, indicating that native-language proficiency accounted for approximately 4.1% of the variance in the combined functional impairment outcomes.

The multivariate effect of country of origin was not statistically significant, Pillai’s Trace = .003, F(2, 314) = 0.55, p = .577, partial η² = .003.

Follow-up univariate analyses and parameter estimates were examined to determine which outcome variables contributed to the significant multivariate effect (Table 11).

**Table 10.** Descriptive Statistics for PTSD and CPTSD Functional Impairment by Home-Language Use (N = 320)

| Home-Language Use | PTSD Functional Impairment (M ± SD) | CPTSD Functional Impairment (M ± SD) |
| --- | --- | --- |
| Never | 3.02 ± 1.40 | 3.50 ± 2.02 |
| Sometimes | 3.20 ± 1.37 | 3.45 ± 2.01 |
| Often | 2.55 ± 1.80 | 3.55 ± 2.00 |
| Always | 3.00 ± 1.50 | 3.71 ± 2.04 |
| Total | 3.07 ± 1.53 | 3.55 ± 2.02 |

*Notes:* PTSD = Post-Traumatic Stress Disorder functional impairment; CPTSD = Complex Post-Traumatic Stress Disorder functional impairment; M = Mean; SD = Standard Deviation. Home-language use reflects frequency of heritage-language use in the household.

**Table 11.**
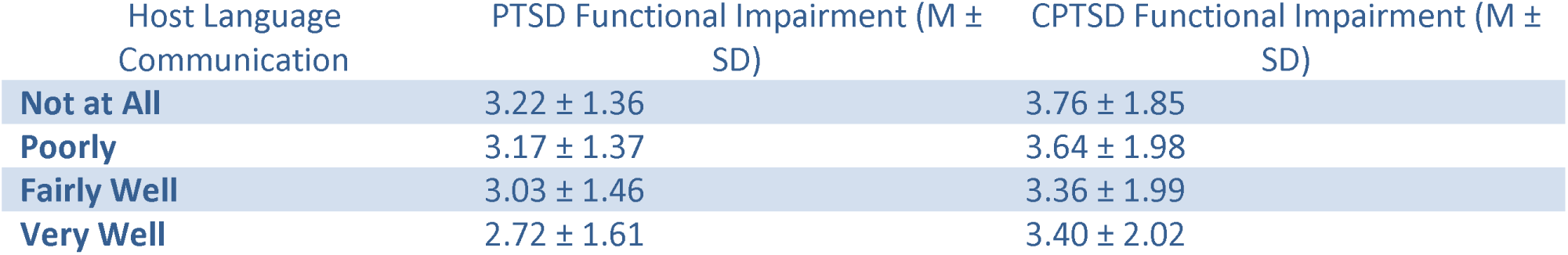
Descriptive Statistics of PTSD and CPTSD Functional Impairment by Host Language Communication (N = 320)

| Host Language Communication | PTSD Functional Impairment (M ± SD) | CPTSD Functional Impairment (M ± SD) |
| --- | --- | --- |
| Not at All | 3.22 ± 1.36 | 3.76 ± 1.85 |
| Poorly | 3.17 ± 1.37 | 3.64 ± 1.98 |
| Fairly Well | 3.03 ± 1.46 | 3.36 ± 1.99 |
| Very Well | 2.72 ± 1.61 | 3.40 ± 2.02 |

**Table 12.** Multivariate Tests of Host Language Communication, Emotional Connection, Country, and Gender on PTSD and CPTSD Functional Impairment.

| Effect | Multivariate Test | F | df1 | df2 | p | Partial $\eta^2$ |
| --- | --- | --- | --- | --- | --- | --- |
| Host Language Communication | Roy's Largest Root | 2.846 | 3 | 311 | 0.038 | 0.027 |
| Emotional Connection to L1 | Pillai's Trace | 1.895 | 6 | 622 | 0.079 | 0.018 |
| Country | Pillai's Trace | 0.750 | 2 | 310 | 0.473 | 0.005 |
| Gender | Pillai's Trace | 1.041 | 2 | 310 | 0.354 | 0.007 |

**Table 13:** Parameter Estimates for Significant Predictors of PTSD Functional Impairment.

| Predictor | Category | B | SE | t | p | Partial $\eta^2$ |
| --- | --- | --- | --- | --- | --- | --- |
| Host Language Communication | Not at All | 0.579 | 0.233 | 2.487 | 0.013 | 0.019 |
| Host Language Communication | Poorly | 0.525 | 0.223 | 2.359 | 0.019 | 0.018 |
| Emotional Connection to L1 | Poorly | -0.482 | 0.238 | -2.027 | 0.044 | 0.013 |

For CPTSD functional impairment, none of the proficiency contrasts differed significantly from the reference category (“very well”). Children who spoke their native language not at all (B = – 0.52, p = .294), poorly (B = 0.19, p = .710), or fairly well (B = 0.58, p = .294) did not differ significantly in CPTSD-related functional impairment. Effect sizes were negligible (partial η² = .000–.004).

For PTSD functional impairment, significant differences were observed across proficiency levels. Compared with children who spoke their native language very well, those who did not speak it at all reported higher PTSD-related functional impairment (B = 1.02, t = 2.73, p = .007, partial η² = .023). Children who spoke their native language poorly (B = 1.23, t = 3.21, p = .001, partial η² = .032) and fairly well (B = 0.94, t = 2.26, p = .025, partial η² = .016) also reported significantly higher PTSD-related functional impairment.

Overall, native-language proficiency was significantly associated with PTSD functional impairment but not with CPTSD functional impairment. Lower levels of native-language proficiency were associated with higher PTSD-related functional impairment, whereas CPTSD-related functional impairment did not differ significantly across proficiency levels.

Figure 1 presents the estimated marginal means of CPTSD functional impairment across levels of native language proficiency. The pattern is nonlinear: impairment increases from “Not at all” (M = 3.15) to a peak at “Fairly well” (M = 4.24), then declines for those who speak their native language very well (M = 3.55). This suggests that partial proficiency may heighten functional impairment, possibly due to identity tension whereas strong proficiency provides cultural stability and reduces trauma-related difficulties.

**Figure 1.**
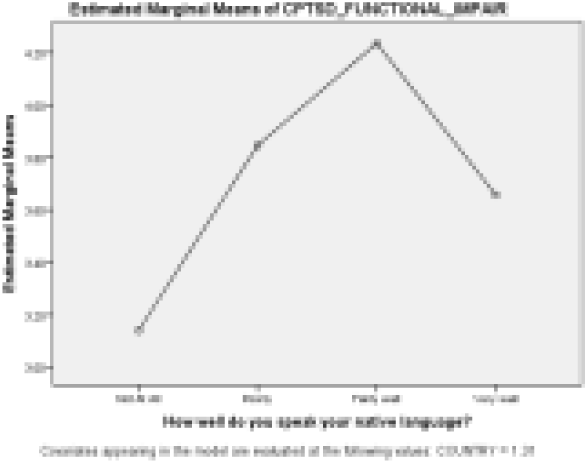
Estimated marginal means of CPTSD functional impairment across levels of native-language proficiency. Note. Figures display estimated marginal means controlling for country (COUNTRY = 1.31). Higher values indicate greater impairment.

Figure 2 shows a nonlinear downward pattern in PTSD functional impairment across levels of native language proficiency. Children who do not speak their native language at all show moderate impairment (≈3.0), but impairment increases slightly for those with poor proficiency (≈3.2), indicating the highest level of PTSD-related functional difficulty. As proficiency improves, impairment begins to decline, dropping to approximately 2.8 among children who speak the language fairly well, and falling sharply to about 2.0 among those who speak it very well. This pattern shows that the relationship is not linear: impairment rises at very low proficiency, then decreases gradually, and finally decreases steeply at high proficiency. Overall, strong native-language proficiency appears to function as a protective factor for PTSD-related functional impairment.

**Figure 2:**
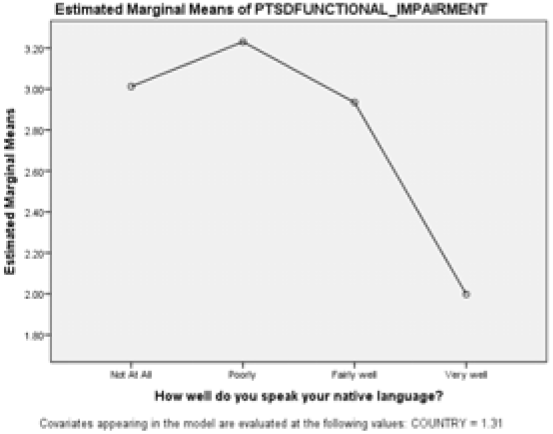
Estimated Marginal Means of PTSD Functional Impairment by Native Language Proficiency. Note. Figures display estimated marginal means controlling for country (COUNTRY = 1.31). Higher values indicate greater impairment.

Hypothesis 2 predicted that patterns of native-language use at home would be associated with trauma-related functional impairment among refugee children. Descriptive statistics for PTSD and CPTSD functional impairment across levels of home-language use are presented in Table 10.

For PTSD, functional impairment was lowest among children whose families often used the native language at home (M = 2.55, SD = 1.80) and highest among those whose families sometimes used it (M = 3.20, SD = 1.37), with intermediate values for the “never” and “always” categories. CPTSD functional impairment showed a less consistent pattern, ranging from M = 3.45 (sometimes) to M = 3.71 (always).

A multivariate analysis of variance (MANOVA) was conducted with PTSD and CPTSD functional impairment as dependent variables and home-language use as the independent variable. Assumptions of homogeneity of covariance matrices and equality of variances were met. The MANOVA revealed a statistically significant multivariate effect of home-language use, Pillai’s Trace = .082, F(6, 630) = 4.50, p < .001, partial η² = .041, indicating that home-language use was associated with variation in trauma-related functional impairment when both outcomes were considered simultaneously.

Follow-up univariate analyses showed a significant main effect of home-language use on PTSD functional impairment, F(3, 316) = 3.50, p = .016, partial η² = .032. In contrast, the univariate effect of home-language use on CPTSD functional impairment was not statistically significant, F(3, 316) = 0.52, p = .670, partial η² = .005.

These results support Hypothesis 2. Home-language use is significantly associated with reduced PTSD-related functional impairment, with frequent (but not exclusive) heritage-language use linked to lower impairment. In contrast, CPTSD functional impairment appears relatively unaffected by home-language practices, reinforcing the conceptual distinction between PTSD and CPTSD and suggesting that ecological and familial language factors are more protective for PTSD than for CPTSD.

#### Hypothesis 3: Host Language Communication, Emotional Connection, and Functional Impairment

Hypothesis 3 examined whether host language communication and emotional connection to language were associated with functional impairment across PTSD and CPTSD outcomes. It was predicted that higher host language communication and stronger emotional connection would correspond to lower functional impairment.

PTSD functional impairment decreased modestly with higher host language proficiency, from 3.22 in the “Not at All” group to 2.72 in the “Very Well” group, suggesting a protective effect of better host language communication. In contrast, CPTSD functional impairment remained relatively stable across groups (3.36–3.76), indicating no clear trend.

#### Multivariate Analysis

A multivariate analysis of variance (MANOVA) was conducted to examine the combined effects of host language communication, emotional connection, country, and gender on PTSD and CPTSD functional impairment. Assumptions of homogeneity of covariance matrices and equality of variances were met.

Host language communication had a small but significant multivariate effect on combined PTSD and CPTSD functional impairment (p = 0.038), while emotional connection showed a non-significant trend (p = 0.079). Country and gender did not significantly affect the combined outcomes.

#### Follow-Up Parameter Estimates

Parameter estimates were examined to clarify the direction and magnitude of significant effects for PTSD functional impairment.

#### Findings

Children reporting “Not at All” or “Poorly” for host language communication had significantly higher PTSD functional impairment compared to the reference category (“Very Well”). Emotional connection to native language showed a small protective effect in the “Poorly” category. No significant predictors emerged for CPTSD functional impairment.

Overall, host language communication has a modest but statistically significant protective effect on PTSD functional impairment, whereas CPTSD functional impairment was largely unaffected by host language communication, emotional connection, country, or gender. Effect sizes were small, indicating these predictors explain only a limited portion of variance.

Conclusively, hypothesis 3 is partially supported: host language communication predicts PTSD outcomes, while emotional connection demonstrates a minor protective effect.

Figure 3 displays estimated marginal means for CPTSD functional impairment across host-language communication levels and emotional connection to language. Although modest descriptive declines in impairment were observed with increasing host-language communication, these trends were shallow and inconsistent across emotional-connection groups. Consistent with univariate analyses, neither host-language communication, emotional connection to native language, nor their interaction significantly predicted CPTSD impairment (all p > .05). Thus, the figure illustrates descriptive variation without statistically supported differences.

**Figure 3:**
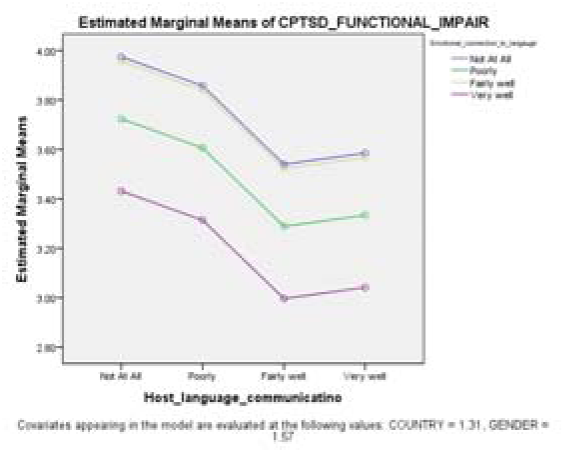
Estimated Marginal Means of CPTSD Functional Impairment Across Levels of Host Language Communication and Emotional Connection to Native Language.

Figure 4 presents the estimated marginal means of PTSD functional impairment across levels of host-language communication and emotional connection to language. PTSD impairment decreases progressively as host-language communication improves across all emotional-connection groups. Estimated means decline from approximately 2.8–3.5 at the “Not at All” communication level to approximately 2.2–2.8 at the “Very Well” level.

**Figure 4:**
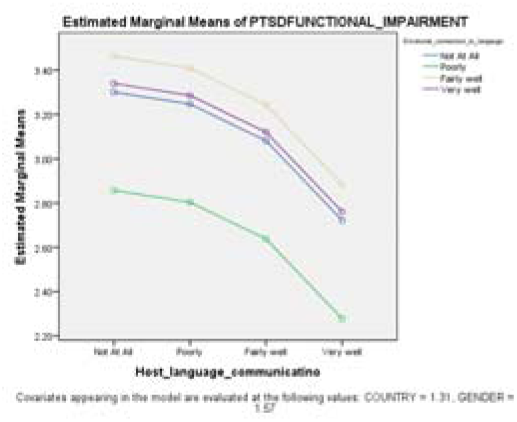
Estimated Marginal Means of PTSD Functional Impairment Across Levels of Host Language Communication and Emotional Connection to Native Language.

Children reporting weaker emotional connection to native language (“Poorly”) demonstrate a descriptively steeper decline (≈ 3.25 to 2.25), whereas those reporting no emotional connection begin at lower impairment levels (≈ 2.85) and decline more gradually. However, these slope differences are descriptive and do not reflect a statistically significant interaction effect.

Consistent with the univariate analyses, host-language communication significantly predicts PTSD functional impairment, whereas emotional connection does not independently predict PTSD outcomes.

#### Hypothesis 4: Heritage-Language Proficiency, Parental/Home-Language Use, Host-Language Use, and Functional Impairment

This hypothesis predicted that heritage-language proficiency, parental/home-language use, and host-language (Nigerian) use will interact to predict PTSD and CPTSD functional impairment among refugee children, such that children with high exposure to both heritage and host languages will exhibit lower functional impairment than children with limited exposure.

#### Descriptive Statistics

Table 14 presents means and standard deviations of CPTSD and PTSD functional impairment across levels of native-language proficiency, parental/home-language use, and Nigerian-language speaking among the sample (N = 320).

**Table 14.** Descriptive Statistics of CPTSD and PTSD Functional Impairment by Language Variables (N = 320)

| Language Variable | Category | N | CPTSD Mean (SD) | PTSD Mean (SD) |
| --- | --- | --- | --- | --- |
| **Native-language proficiency** | Not at all | 96 | 3.16 (2.13) | 3.00 (1.47) |
|  | Poorly | 95 | 3.84 (1.75) | 3.23 (1.27) |
|  | Fairly well | 45 | 4.24 (1.32) | 2.93 (1.54) |
|  | Very well | 17 | 3.65 (2.15) | 2.00 (1.84) |
| **Parents/grandparents speak dialect at home** | Never | 187 | 3.53 (1.94) | 3.02 (1.44) |
|  | Sometimes | 71 | 3.45 (2.01) | 3.20 (1.37) |
|  | Often | 31 | 3.55 (2.03) | 2.55 (1.80) |
|  | Always | 31 | 3.71 (2.04) | 3.00 (1.51) |
| **Child speaks a Nigerian language** | Yes | 251 | 3.50 (1.97) | 2.97 (1.50) |
|  | No | 67 | 3.69 (1.95) | 3.13 (1.39) |

Table 14 presents PTSD functional impairment generally decreases with higher native-language proficiency, with the lowest PTSD mean observed for children reporting “Very well” (M = 2.00). CPTSD patterns are less linear, showing modest increases at higher proficiency levels. Children from homes where dialects are spoken more frequently or who speak a Nigerian language tend to show slightly lower PTSD and CPTSD impairment. Overall, these descriptive patterns suggest that both heritage-language competence and regular exposure to home or host languages may provide modest protective effects against trauma-related functional impairment.

#### Multivariate Analysis

A MANOVA was conducted to examine the main effects and interactions of native-language proficiency, parental/home-language use, and Nigerian-language speaking on PTSD and CPTSD functional impairment.

Table 15 shows that **s**ignificant main effects were observed for native-language proficiency on PTSD (F(3, 290) = 3.44, p = .017, partial η² = .034) and nation considered home (F(1, 290) = 6.07, p = .014, partial η² = .020). A significant two-way interaction between parental language use and Nigerian-language speaking emerged for PTSD (F(3, 290) = 3.50, p = .016, partial η² = .035). For CPTSD, the interaction between native-language proficiency and parental language use approached significance (F(9, 290) = 1.92, p = .049, partial η² = .056). These findings indicate that both heritage and host-language environments jointly influence PTSD functional impairment more consistently than CPTSD.

**Table 15.**
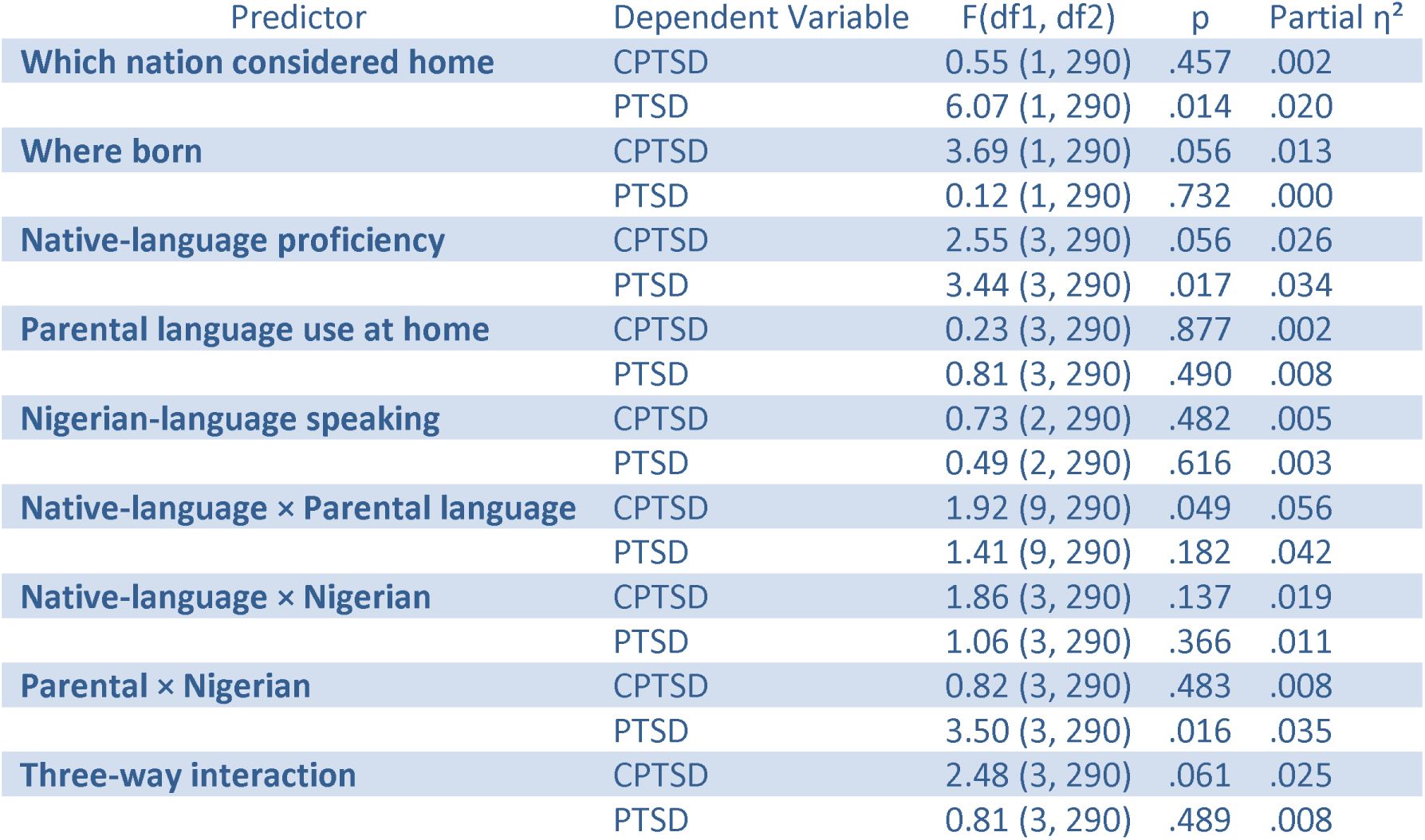
MANOVA: Tests of Between-Subjects Effects on PTSD and CPTSD Functional Impairment.

#### Estimated Marginal Means / Three-Way Interaction

As seems in Table 16**, PTSD f**unctional impairment decreases consistently as native-language proficiency increases, with the highest PTSD impairment at “Not at All” (M = 3.18) and the lowest at “Very well” (M = 1.86).**CPTSD functional i**mpairment shows a less linear trend, with the highest impairment at the “Very well” proficiency level (M = 4.12). Three-way interaction: Linear regression revealed that children with low native-language proficiency, low parental/home-language use, and non-use of Nigerian language had significantly higher CPTSD functional impairment (B = 6.09, p = .012, 95% CI [1.32, 10.85]). No other main effects or interactions were statistically significant.

**Table 16.** Estimated Marginal Means of CPTSD and PTSD Functional Impairment Across Native-Language Proficiency.

| Native-Language Proficiency | CPTSD M (SE) | 95% CI | PTSD M (SE) | 95% CI |
| --- | --- | --- | --- | --- |
| **Not at All** | 2.64 (0.31) | [2.03, 3.26] | 3.18 (0.24) | [2.71, 3.64] |
| **Poorly** | 3.64 (0.28) | [3.09, 4.18] | 3.12 (0.21) | [2.71, 3.53] |
| **Fairly well** | 3.97 (0.43) | [3.11, 4.82] | 2.62 (0.33) | [1.97, 3.27] |
| **Very well** | 4.12 (0.61) | [2.91, 5.33] | 1.86 (0.47) | [0.94, 2.77] |

**Table 21.** Descriptive Statistics of CPTSD and PTSD Functional Impairment by Trauma Type and Exposure Level (Collapsed)

| Trauma Type | Exposure Level | N | CPTSD Functional Impairment Mean (SD) | PTSD Functional Impairment Mean (SD) |
| --- | --- | --- | --- | --- |
| **Witnessed Trauma** | Low (0–2) | 148 | 3.30 (1.97) | 3.42 (2.07) |
|  | Moderate (3–4) | 82 | 4.24 (1.20) | 4.28 (1.11) |
|  | High (5–6) | 90 | 3.85 (1.82) | 4.30 (0.89) |
| **Physical Trauma** | Low (0–2) | 175 | 3.20 (2.04) | 3.52 (1.84) |
|  | Moderate (3–4) | 55 | 3.28 (2.15) | 3.47 (1.66) |
|  | High (5) | 90 | 4.00 (1.64) | 4.24 (1.59) |

**Table 23.** Tests of Between-Subjects Effects.

| Source | Dependent Variable | F | df | Sig. | Partial $\eta^2$ |
| --- | --- | --- | --- | --- | --- |
| **Witnessed Trauma** | CPTSD | 9.201 | 6, 305 | < .001 | 0.153 |
| **Physical Trauma** | CPTSD | 1.400 | 5, 305 | .224 | 0.022 |
| **Witnessed Trauma** | PTSD | 4.160 | 6, 305 | .042 | 0.076 |
| **Physical Trauma** | PTSD | 2.747 | 5, 305 | .099 | 0.043 |
| **Gender** | CPTSD | 0.260 | 1, 305 | .610 | 0.001 |
| **Gender** | PTSD | 1.665 | 1, 305 | .198 | 0.005 |
| **Country** | CPTSD | 2.102 | 1, 305 | .148 | 0.007 |
| **Country** | **PTSD** | 0.383 | 1, 305 | .537 | 0.001 |
| **Most Bothersome** | **CPTSD** | 0.039 | 1, 305 | .844 | 0.000 |
| **Most Bothersome** | **PTSD** | 2.718 | 1, 305 | .100 | 0.009 |

**Table 24.** Parameter Estimates for CPTSD and PTSD Functional Impairment.

| **Dependent Variable** | **Predictor** | **B** | **SE** | **t** | **p** | **95% CI** | **Partial $\eta^2$** |
| --- | --- | --- | --- | --- | --- | --- | --- |
| **CPTSD** | Intercept | 4.414 | 0.539 | 8.194 | < .001 | 3.354, 5.474 | 0.180 |
| **CPTSD** | Witnessed Trauma = 0 | -1.654 | 0.308 | -5.370 | < .001 | -2.260, -1.048 | 0.086 |
| **CPTSD** | Physical Trauma = 1 | -0.826 | 0.363 | -2.273 | .024 | -1.540, -0.111 | 0.017 |
| **CPTSD** | Gender | 0.108 | 0.211 | 0.510 | .610 | -0.308, 0.523 | 0.001 |
| **CPTSD** | Country | -0.329 | 0.227 | -1.450 | .148 | -0.776, 0.118 | 0.007 |
| **PTSD** | Intercept | 4.785 | 0.510 | 9.378 | < .001 | 3.781, 5.789 | 0.224 |
| **PTSD** | Witnessed Trauma = 0 | -0.817 | 0.292 | -2.801 | .005 | -1.391, -0.243 | 0.025 |
| **PTSD** | Physical Trauma = 2 | -0.879 | 0.344 | -2.555 | .011 | -1.556, -0.202 | 0.021 |
| **PTSD** | Gender | −0.258 | 0.200 | −1.290 | .198 | −0.652,<br>0.136 | 0.005 |
| **PTSD** | Country | 0.133 | 0.215 | 0.619 | .537 | −0.290,<br>0.556 | 0.001 |

In summary, **t**hese results provide partial support for Hypothesis 4: combined heritage and host-language exposure is associated with lower trauma-related functional impairment, particularly for children at high risk for CPTSD. The findings highlight the importance of multi-level language environments in shaping refugee children’s psychological outcomes.

Figure 5 shows the estimated marginal means of CPTSD functional impairment across native-language proficiency by Nigerian-language speaking status. Among children who did not speak a Nigerian language, CPTSD impairment increased descriptively with higher native-language proficiency, rising from approximately 2.4 at “Not at all” to about 5.0 at “Very well.” In contrast, among Nigerian-language speakers, CPTSD impairment remained relatively stable across proficiency levels (approximately 3.3 to 3.9). The divergence between groups was most evident at higher proficiency levels, where non–Nigerian-language speakers exhibited roughly 1.1 points higher CPTSD impairment. This pattern suggests that increases in native-language proficiency are associated with higher CPTSD impairment primarily among children who do not use a Nigerian language; however, these trends are descriptive and should be interpreted alongside the corresponding statistical tests.

**Figure 5.**
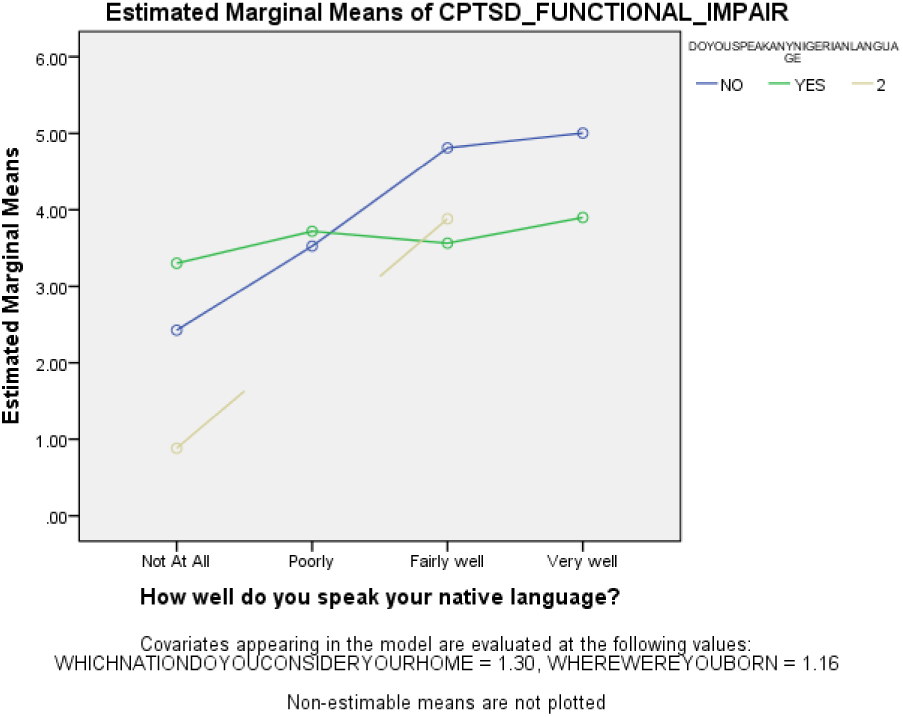
Estimated marginal means interaction plot showing CPTSD functional impairment across levels of native-language proficiency by Nigerian-language use.

Figure 6 presents estimated marginal means of PTSD functional impairment across native-language proficiency levels by Nigerian-language use. The Native-language × Nigerian-language interaction was not statistically significant, F(3, 290) = 1.06, p = .366. Nonetheless, descriptive trends differed across groups. Among Nigerian-language speakers, impairment declined progressively from approximately 3.0 at low proficiency to 1.7 at high proficiency. Among non–Nigerian-language speakers, impairment decreased at intermediate proficiency (≈1.6) but increased again at the highest level (≈2.4), indicating a less consistent pattern. Although not statistically confirmed, these trends suggest that PTSD functional improvement associated with higher native-language proficiency may be more stable among children who engage in the host language.

**Figure 6.**
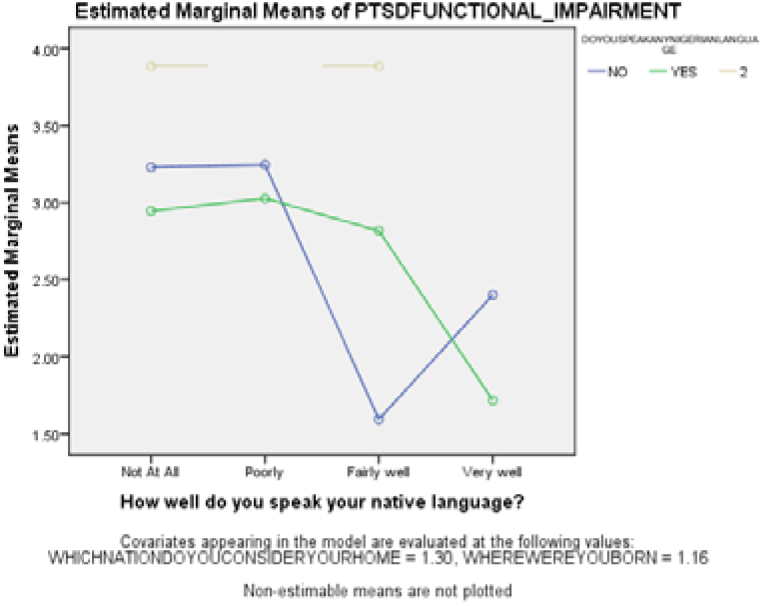
Estimated marginal means interaction plot showing PTSD functional impairment across levels of native-language proficiency by Nigerian-language use.

**Figure 7.**
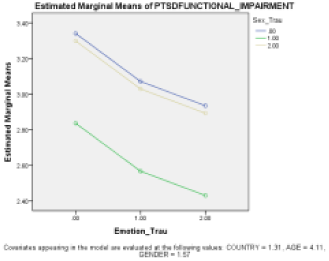
Estimated Marginal Means of PTSD Functional Impairment Across Levels of Emotional Connection to Language by Host-Language Communication.

**Figure 8.**
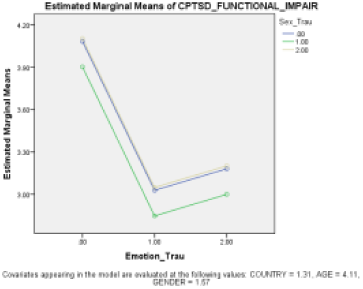
Estimated Marginal Means of CPTSD Functional Impairment Across Levels of Emotional Connection to Language by Sex.

Hypothesis 5: Emotional and sexual trauma exposure will be significantly associated with variation in PTSD and CPTSD functional impairment in refugee children, with trauma exposure expected to relate to differences in impairment levels.

Descriptive analyses indicated that PTSD functional impairment varied modestly across levels of emotional trauma, increasing slightly from M = 3.42 (SD = 1.88) among children reporting no emotional trauma to M = 3.78 (SD = 1.94) at high emotional trauma. PTSD impairment remained largely stable across sexual trauma levels, with means ranging from 3.56 to 3.63. In contrast, CPTSD functional impairment demonstrated greater variation across emotional trauma exposure. The highest impairment was observed among children reporting no emotional trauma (M = 4.01, SD = 1.95), followed by moderate (M = 2.95, SD = 1.87) and high (M = 3.11, SD = 1.90) levels, reflecting a non-linear pattern. CPTSD impairment varied minimally across sexual trauma levels, ranging from 3.29 to 3.38. These patterns indicate that CPTSD and PTSD functional impairment responded differently to trauma exposure in this sample.

Multivariate analysis of variance (MANOVA) confirmed these differences. Emotional trauma significantly predicted overall functional impairment, Wilks’ Λ = .947, F(4, 620) = 4.41, p = .002, partial η² = .028, whereas sexual trauma also had a smaller but significant effect, Wilks’ Λ = .961, F(4, 620) = 3.10, p = .015, partial η² = .019. The interaction between emotional and sexual trauma was significant, Wilks’ Λ = .969, F(8, 620) = 2.42, p = .014, partial η² = .020, indicating that the effect of trauma exposure on functional impairment is dependent on the combination of trauma types.

Univariate tests further clarified these effects. PTSD functional impairment was not significantly predicted by either emotional trauma (F(2, 312) = 2.63, p = .074) or sexual trauma (F(2, 312) = 2.68, p = .070). In contrast, CPTSD functional impairment was significantly predicted by emotional trauma (F(2, 312) = 8.69, p < .001, partial η² = .053). Although sexual trauma alone did not have a main effect, the interaction of emotional and sexual trauma was significant for CPTSD outcomes (p = .024, partial η² = .016), highlighting differential sensitivity to trauma type and trauma interactions.

Overall, Hypothesis 5 received partial support. Emotional trauma significantly predicted CPTSD functional impairment, whereas PTSD functional impairment showed minimal variation across trauma levels. Sexual trauma did not independently predict impairment but moderated CPTSD outcomes, demonstrating that CPTSD and PTSD functional impairment operate differently in response to trauma exposure in refugee children.

The slope plot for PTSD functional impairment across emotional trauma levels shows a slight upward trend, from M = 3.42 (none) to M = 3.78 (high), consistent with descriptive statistics. Lines representing sexual trauma levels indicate a modest interaction: PTSD impairment is slightly higher at lower emotional trauma when sexual trauma is absent and slightly lower when sexual trauma is higher. These visual patterns align with the statistical analyses. Emotional trauma alone did not significantly predict PTSD impairment (F(2, 312) = 2.63, p = .074, partial η² = .016), whereas the emotional × sexual trauma interaction was significant in the multivariate model (Wilks’ Λ = .969, F(8, 620) = 2.42, p = .014, partial η² = .020).

As illustrated in Figure X, CPTSD-related functional impairment declines sharply as Emotional trauma increases from 0 to 1, with estimated slopes ranging from approximately −2.0 to −2.2 across exposure levels. This marked reduction suggests that initial increases in emotional engagement are associated with substantial improvements in daily functioning among refugee children. Consistent with trauma models of CPTSD, early shifts in emotional processing may alleviate avoidance and numbing responses that constrain adaptive functioning.

Notably, from emotional trauma levels 1 to 2, the slope becomes modestly positive (approximately +0.1 to +0.5), indicating a slight increase in functional impairment at higher levels of emotional engagement. This nonlinear trend aligns with trauma-informed frameworks suggesting that deeper emotional processing can temporarily intensify distress as previously suppressed trauma-related affect becomes more accessible. Together, these findings suggest that emotional engagement operates as a complex mechanism in the association between trauma exposure and CPTSD-related impairment, with early gains yielding substantial functional improvements and later increases potentially reflecting transitional adjustment processes.

Hypothesis 6 states that trauma exposure—specifically witnessed trauma and physical trauma—would significantly predict functional impairment associated with CPTSD and PTSD among refugee children, such that higher levels of trauma exposure would be associated with greater impairment

#### Descriptive Statistics

Descriptive statistics indicated that functional impairment generally increased with greater trauma exposure, although the pattern was not strictly linear. For witnessed trauma, children with low exposure reported the lowest levels of functional impairment for both CPTSD (M = 3.30, SD = 1.97) and PTSD (M = 3.42, SD = 2.07). Functional impairment was highest at moderate exposure levels for CPTSD (M = 4.24, SD = 1.20) and PTSD (M = 4.28, SD = 1.11), while high exposure levels remained elevated but slightly lower for CPTSD, indicating variability in impairment levels among children with extensive exposure.

For physical trauma, functional impairment was lowest at low exposure levels and increased with severity, with the highest impairment observed at high exposure for both CPTSD (M = 4.00, SD = 1.64) and PTSD (M = 4.24, SD = 1.59). Overall, these descriptive patterns are consistent with the expectation that greater trauma exposure is associated with higher levels of functional impairment, while also suggesting heterogeneity in symptom expression across exposure levels and trauma types.

### Multivariate Effects

#### Tests (MANOVA) for CPTSD and PTSD Functional Impairment

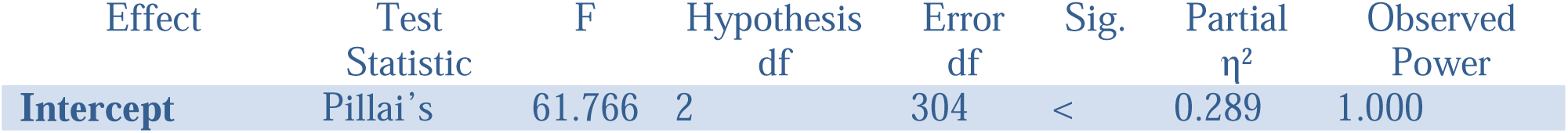
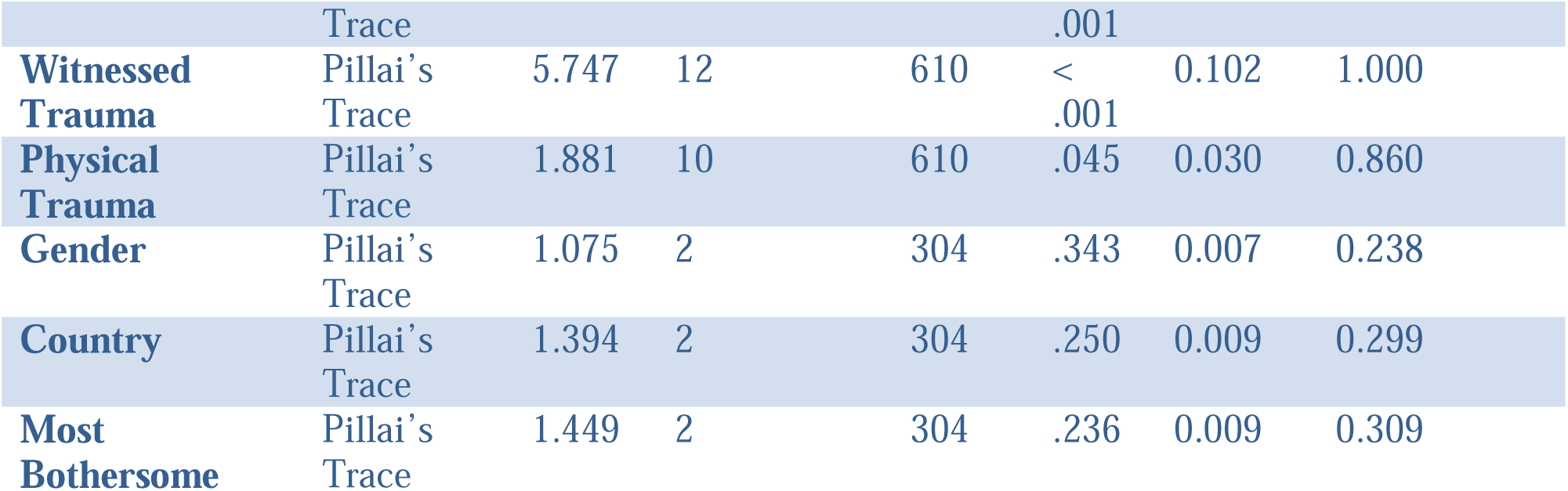

Multivariate analyses using Pillai’s Trace revealed a statistically significant effect of witnessed trauma on the combined CPTSD and PTSD functional impairment outcomes, *F*(12, 610) = 5.747, *p* < .001, with a medium multivariate effect size (partial η² = .102). Physical trauma also demonstrated a statistically significant multivariate effect, *F*(10, 610) = 1.881, *p* = .045, although the associated effect size was small (partial η² = .030). In contrast, gender, country of origin, and the most bothersome trauma-related factor did not significantly predict the combined functional impairment outcomes (all *p* > .05).

These findings indicate that trauma exposure—particularly witnessed trauma—plays a central role in explaining variation in functional impairment among refugee children when CPTSD and PTSD outcomes are considered jointly.

#### Univariate Effects

Univariate analyses demonstrated that witnessed trauma was a significant predictor of CPTSD functional impairment, *F*(6, 305) = 9.201, *p* < .001, partial η² = .153, and PTSD functional impairment, *F*(6, 305) = 4.160, *p* = .042, partial η² = .076. The magnitude of the effect was larger for CPTSD than for PTSD, indicating greater sensitivity of CPTSD-related functional impairment to variations in witnessed trauma exposure.

Physical trauma was not a significant predictor of CPTSD functional impairment, *F*(5, 305) = 1.400, *p* = .224, and showed a trend-level association with PTSD functional impairment, *F*(5, 305) = 2.747, *p* = .099. Gender, country of origin, and the most bothersome trauma-related factor were not significant predictors of functional impairment for either CPTSD or PTSD.

#### Parameter Estimates

Parameter estimates indicated that children with no exposure to witnessed trauma reported significantly lower functional impairment for both CPTSD (B = −1.654, *p* < .001) and PTSD (B = −0.817, *p* = .005) compared to exposed children. Physical trauma was also associated with lower functional impairment for PTSD (B = −0.879, *p* = .011) and CPTSD (B = −0.826, *p* = .024), although the corresponding effect sizes were small. Gender and country of origin were not significant predictors in either model.

The emergence of physical trauma effects in the parameter estimates suggests that, when adjusting for witnessed trauma and other covariates, physical trauma contributes independently—though modestly—to functional impairment outcomes.

#### Overall Summary of Findings

Collectively, these results support Hypothesis 6. Witnessed trauma emerged as the most robust trauma-related predictor of functional impairment, particularly for CPTSD, while physical trauma demonstrated smaller and more variable associations, most evident for PTSD. Demographic factors showed minimal influence across models, underscoring trauma exposure—especially witnessed trauma—as a central determinant of functional impairment among refugee children.

Figure 9 illustrates the estimated marginal means of CPTSD functional impairment across increasing levels of witnessed trauma, stratified by physical trauma exposure, with covariates held constant. Across all levels of physical trauma, CPTSD functional impairment rises sharply from no witnessed trauma to initial exposure. Specifically, at witnessed trauma = 0, impairment ranges from approximately 1.7 to 2.6 across physical trauma categories, but increases substantially at witnessed trauma = 1 to approximately 3.5–4.3, representing an increase of nearly two points. Thereafter, impairment remains elevated, peaking around witnessed trauma levels 3–4 with estimated means between 3.7 and 4.6, and stabilizing or slightly declining at the highest exposure levels (5–6), where values remain high (approximately 3.3–4.2). This pattern closely reflects the strong univariate effect of witnessed trauma on CPTSD functional impairment (F = 9.201, p < .001, partial η² = .153), indicating a substantial and clinically meaningful association. Physical trauma contributes a secondary, additive effect, consistently shifting impairment upward across all levels of witnessed trauma, though the magnitude of this shift is modest. For example, at witnessed trauma = 3, impairment increases from approximately 3.7 among children with no physical trauma to about 4.5 among those with the highest physical trauma exposure, a difference of less than one point; a similar pattern is observed at witnessed trauma = 4. The near-parallel trajectories across physical trauma levels indicate that physical trauma does not alter the slope of the witnessed trauma–impairment relationship, consistent with its non-significant univariate effect and small multivariate effect size (partial η² = .030). Overall, the figure demonstrates that witnessed trauma is the dominant predictor of CPTSD functional impairment, producing large increases in impairment across exposure levels, while physical trauma adds a smaller, additive elevation without evidence of interaction.

**Figure 9.**
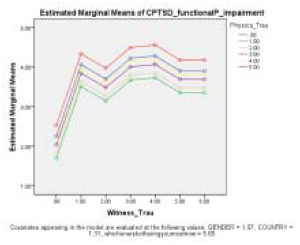
Estimated Marginal Means of CPTSD Functional Impairment by Witnessed Trauma and Physical Trauma.

Figure 9 illustrates the estimated marginal means of CPTSD functional impairment across increasing levels of witnessed trauma, stratified by physical trauma exposure, with covariates held constant. Across all levels of physical trauma, CPTSD functional impairment rises sharply from no witnessed trauma to initial exposure. Specifically, at witnessed trauma = 0, impairment ranges from approximately 1.7 to 2.6 across physical trauma categories, but increases substantially at witnessed trauma = 1 to approximately 3.5–4.3, representing an increase of nearly two points. Thereafter, impairment remains elevated, peaking around witnessed trauma levels 3–4 with estimated means between 3.7 and 4.6, and stabilizing or slightly declining at the highest exposure levels (5–6), where values remain high (approximately 3.3–4.2). This pattern closely reflects the strong univariate effect of witnessed trauma on CPTSD functional impairment (F = 9.201, p < .001, partial η² = .153), indicating a substantial and clinically meaningful association. Physical trauma contributes a secondary, additive effect, consistently shifting impairment upward across all levels of witnessed trauma, though the magnitude of this shift is modest. For example, at witnessed trauma = 3, impairment increases from approximately 3.7 among children with no physical trauma to about 4.5 among those with the highest physical trauma exposure, a difference of less than one point; a similar pattern is observed at witnessed trauma = 4. The near-parallel trajectories across physical trauma levels indicate that physical trauma does not alter the slope of the witnessed trauma–impairment relationship, consistent with its non-significant univariate effect and small multivariate effect size (partial η² = .030). Overall, the figure demonstrates that witnessed trauma is the dominant predictor of CPTSD functional impairment, producing large increases in impairment across exposure levels, while physical trauma adds a smaller, additive elevation without evidence of interaction.

## Discussion

This study explored the sociolinguistic–psychological dynamics linking heritage-language proficiency, home-language practices, host-language engagement, and trauma exposure to PTSD and CPTSD-related functional impairment among Liberian and Sierra Leonean refugee children in Nigeria. The findings indicate that language competence and patterns of language use are not neutral background variables but are meaningfully associated with trauma-related functional outcomes. In particular, reduced heritage-language proficiency and shifting home-language practices appear to intersect with traumatic exposure in shaping levels of impairment, lending empirical support to concerns about language attrition and semilingualism in displacement contexts. The absence of strong demographic effects further underscores that linguistic dislocation, rather than static background characteristics, may constitute a central mechanism through which trauma impacts everyday functioning. These results advance a contextualized understanding of mental health that integrates linguistic identity, emotional expression, and structural vulnerability within stateless refugee communities.

Hypothesis 1 posited that native-language proficiency would inversely predict PTSD- and CPTSD-related functional impairment among stateless Liberian and Sierra Leonean refugee children. Consistent with the hypothesis, PTSD functional impairment was significantly moderated by native-language competence. Children with minimal or poor proficiency exhibited elevated impairment, whereas those with strong proficiency demonstrated a marked reduction in PTSD-related functional difficulties. Estimated marginal means (Figure 2) revealed a nonlinear, dose-dependent protective trajectory, indicating that high native-language competence functions as a psychosocial buffer by supporting emotional articulation, culturally embedded coping strategies, and relational attunement within the family microsystem.

For CPTSD functional impairment, slopes (Figure 1) displayed an inverted-U pattern, with children demonstrating partial native-language proficiency experiencing the highest functional disruption, while very low or very high proficiency corresponded with relatively lower impairment. This suggests that partial language competence may generate identity tension, reflecting ambivalent integration of cultural and emotional schemas, whereas robust proficiency restores cultural scaffolding, which can mitigate complex trauma sequelae to some extent. Collectively, these patterns underscore the nonlinear and outcome-specific protective effects of native-language competence, highlighting differential mechanisms for discrete versus complex trauma outcomes.

By situating these findings within the structural precarity and prolonged statelessness of Liberian and Sierra Leonean refugee children in Nigeria, the study also illustrates diagnostic differentiation between PTSD and CPTSD in a West African pediatric context. PTSD, according to ICD-11, reflects discrete, event-specific trauma reactions and related functional impairment, whereas CPTSD captures chronic, cumulative trauma effects, including disturbances in self-organization (affect dysregulation, negative self-concept, and relational dysfunction). The observed protective effect of native-language proficiency for PTSD—but its limited impact on CPTSD—reflects these distinctions: linguistic continuity mitigates acute, event-specific stress, but cannot fully compensate for systemic, multilevel deficits (e.g., disrupted attachment, intergenerational marginalization, and structural statelessness) that drive CPTSD.

Theoretically, these findings extend Ethnolinguistic Identity Theory (Tajfel & Turner, 1979) by demonstrating that language continuity underpins identity stability and functional adaptation, while partial erosion amplifies vulnerability under chronic adversity. Empirically, the study advances the literature by quantifying nonlinear, outcome-specific effects of native-language competence on trauma-related functional impairment among stateless refugee children—a demographic rarely examined in West African contexts. These results also align with UNESCO’s (2020) advocacy for mother-tongue multilingual education and Becker and Magno’s (2022) emphasis on inclusive language policies as psychosocial scaffolds, while illustrating that native-language continuity alone may be insufficient to attenuate complex, cumulative trauma outcomes, highlighting the need for multilevel, context-sensitive interventions targeting CPTSD. Hypothesis 2 examined whether patterns of native-language use in the home are associated with reduced PTSD functional impairment but not CPTSD functional impairment. The results demonstrate a significant association, suggesting that home-language practices interact with trauma-related cognitive constraints to influence functional outcomes.

Extensive research on the Home Language and Literacy Environment (HLLE) shows that consistent exposure to a native language supports cognitive, linguistic, and academic development (Vygotsky, 1978; Weldemariam, 2022; Yeomans-Maldonado, 2021). In bilingual or multilingual households, structured and intentional language use promotes executive functioning and cognitive flexibility (Filippi et al., 2022; Planckaert, 2023; Tran et al., 2022). However, these benefits are primarily documented in stable developmental contexts and have rarely been explored among trauma-exposed or displaced populations.

Cognitive models of PTSD suggest that trauma exposure may be associated with reductions in working memory capacity (WMC), which can affect language production, emotional regulation, and everyday functioning (Norte et al., 2024). Working memory plays a central role in linguistic processing, including lexical retrieval and syntactic planning (Badecker & Kuminiak, 2007; Deldar et al., 2020), and limitations in this system have been linked to communication difficulties in both clinical and developmental populations (Kolk, 1995; Boye et al., 2023; Fedorenko et al., 2023). Although working memory and executive functioning were not directly assessed in the present study, the observed association between inconsistent home-language use and functional impairment may be interpreted in light of cognitive models of PTSD and working-memory accounts of language processing, which suggest that trauma-related cognitive constraints can increase the effort required to manage multiple linguistic systems. In contexts where children must alternate between languages or operate with unstable linguistic input, the resulting cognitive load may exceed available regulatory resources, potentially contributing to greater difficulty in communication, emotional control, and daily functioning.

Cognitive models of PTSD suggest persistent reductions in working memory capacity (WMC), which impair language production, emotional regulation, and daily functioning (Norte et al., 2024). Working memory supports key linguistic processes, including lexical retrieval and syntactic planning (Badecker & Kuminiak, 2007; Deldar et al., 2020). Impairments in WMC, similar to those observed in conditions like Broca’s aphasia, may underlie difficulties in everyday communication (Kolk, 1995; Boye et al., 2023; Fedorenko et al., 2023). Trauma-related cognitive constraints thus overlap with mechanisms critical for managing multiple languages.

The observed non-linear association—with the highest functional impairment among children from households reporting inconsistent (“sometimes”) native-language use—can be interpreted in light of cognitive accounts of bilingual processing and trauma-related cognitive constraints. Managing multiple languages requires substantial regulatory resources, particularly for switching between languages and inhibiting competing systems. Cognitive models of PTSD suggest that trauma exposure may be associated with reductions in working memory capacity, a function that supports both language control and emotional regulation. In this context, inconsistent home-language use may place additional cognitive demands on children who already experience reduced regulatory capacity, potentially contributing to greater difficulty in communication, emotional control, and daily functioning. Thus, inconsistent language environments may reflect not only linguistic variation but situations in which cognitive load approaches or exceeds available resources.

These findings extend HLLE research by highlighting boundary conditions: while consistent home-language use supports cognitive and social development in stable contexts, variability in language exposure may signal broader family stress and environmental unpredictability under trauma and displacement. Consistent with prior evidence that trauma can alter language production and fluency (Isen & Boye, 2023), our results suggest that home-language use functions as both a protective and regulatory factor, supporting expressive language capacity, emotional regulation, and social engagement in refugee children (Hamuddin, Ramadhani, & Ningrum, 2024).

In sum, Hypothesis 2 is supported: patterns of native-language use at home are significantly associated with functional impairment among refugee children. These results are best understood through an integrated HLLE–cognitive load perspective, in which trauma-related working memory reductions interact with home communication patterns to shape functional outcomes. Future research incorporating direct measures of working memory, caregiver mental health, and language-switching frequency would further clarify these mechanisms.

Hypothesis 3 examined whether host-language communication and emotional connection to parental language (L1) were associated with functional impairment across PTSD and CPTSD outcomes. Descriptive analyses indicated a monotonic decline in PTSD functional impairment as host-language communication increased, from M = 3.22 (“Not at All”) to M = 2.72 (“Very Well”), suggesting a protective effect of host-language proficiency. CPTSD functional impairment, by contrast, remained largely invariant across host-language levels (M = 3.36–3.76), reflecting the entrenched, multi-systemic nature of CPTSD. Multivariate analyses corroborated these patterns: host-language communication exerted a small but significant multivariate effect (Roy’s Largest Root = 2.846, p = 0.038, partial η² = 0.027), whereas emotional connection to L1 exhibited a non-significant trend (Pillai’s Trace = 1.895, p = 0.079). Country of origin and gender did not significantly predict functional impairment.

Figures 3 and 4 provide visual corroboration. Figure 4 demonstrates that PTSD functional impairment decreases progressively with increasing host-language communication, independent of emotional connection to L1, whereas Figure 3 illustrates that CPTSD functional impairment remains largely stable across both linguistic variables. These figures reinforce the analytic conclusion that host-language proficiency mitigates PTSD-related functional impairment, whereas CPTSD deficits are resistant to single-domain linguistic resources.

From an Ecological Systems Theory (EST) perspective (Bronfenbrenner, 1977; Shelton, 2019), host-language proficiency facilitates adaptive engagement within nested microsystems (familial and peer networks) and mesosystems (schools, community institutions), enabling participation, social integration, and functional coping that attenuate PTSD impairment. CPTSD, characterized by persistent dysregulation of affect, self-concept, and relational capacities, is predominantly shaped by exosystemic and macrosystemic disruptions, including restricted access to institutional support, sociopolitical marginalization, and post-conflict socio-economic precarity (Anderson et al., 2004; Peterson & Bush, 2013). Chronosystemic dynamics, such as the cessation of UNHCR services, amplify vulnerability and prolong functional deficits (Anderson et al., 2004; Betancourt et al., 2022).

Drawing on bilingualism research, early claims that bilingualism impedes cognitive development (Smith, 1939; Brumberg, 1986; Hakuta & Diaz, 1985) have been superseded by evidence demonstrating that fluent bilingual children exhibit enhanced cognitive flexibility, metalinguistic awareness, and academic achievement (Peal & Lambert, 1962; Leopold & Saporta, 1961; Cummins, 1978; Rumbaut, 1995; Portes & Rumbaut, 1996). Among Liberian and Sierra Leonean refugee children in Oru Camp, semi-lingualism emerges from structural constraints—including limited parental L1 fluency, English-only schooling, and socialization in local Nigerian languages—underscoring the functional salience of host-language proficiency for mitigating PTSD-related functional impairment.

Moreover, language choice is modulated by affective salience and sociolinguistic context. Multilingual individuals often select languages congruent with the emotional valence of the situation: emotionally intimate contexts—such as expressions of love, grief, or anger—are preferentially mediated in L1 due to its deep affective resonance, whereas socially evaluative or public contexts invoke host or community languages perceived as neutral or instrumental (Schwieter & Sunderman, 2020; Fatima et al., 2024). Among Oru Camp refugee children, host languages such as Yoruba or Igbo may serve instrumental functions, but intergenerational heritage-language attrition limits opportunities to develop emotional attachment to L1, with downstream implications for socio-emotional expression and identity consolidation.

These findings have theoretical and empirical contributions. These findings delineate a functional dissociation between PTSD and CPTSD: host-language communication is significantly associated with reduced PTSD functional impairment, whereas CPTSD functional deficits remain largely refractory to linguistic resources. Emotional connection to L1, while theoretically pertinent per Ethnolinguistic Identity Theory (Tajfel & Turner, 1979), did not significantly mitigate functional impairment in this cohort, highlighting the need for future studies to examine whether more intensive heritage-language engagement could buffer CPTSD outcomes. Collectively, the study advances the literature by integrating linguistic, ecological, and trauma-informed perspectives, illustrating how host-language proficiency and heritage-language dynamics interface with functional adaptation in refugee children.

Hypothesis 4 investigated the complex interplay between children’s heritage-language (HL) proficiency, parental/home-language use, and host-language engagement in predicting PTSD and CPTSD functional impairment, positing that multilayered linguistic scaffolding would attenuate trauma-related dysfunction. Analyses incorporated both two-way and three-way interaction models to elucidate configuration-dependent linguistic effects, consistent with ecological and family-systems perspectives on trauma resilience.

Indeed, the two-way interaction between parental/home-language use and host-language proficiency was statistically significant, F(3, 290) = 3.50, p = .016, partial η² = .035, indicating that the salutary effects of host-language competence on PTSD-related functional impairment were contingent upon active heritage-language support in the home environment. This synergistic effect is visually depicted in Figure 6. For Nigerian-language speakers, PTSD impairment exhibited a monotonic decrement with increasing HL proficiency (≈3.0 → 1.7), reflecting robust protective synergy. Conversely, non–Nigerian-language speakers displayed a non-linear trajectory: impairment decreased at intermediate HL proficiency (≈1.6) but resurged at maximal proficiency (≈2.4), suggesting that HL engagement alone may not confer uniform protection absent culturally congruent home-language reinforcement.

These findings underscore the conditional nature of linguistic resilience in PTSD outcomes. They align with extant scholarship emphasizing the psychosocial utility of heritage-language maintenance within familial contexts. For example, Kang (2013) demonstrates that HL use reinforces ethnic identity and familial cohesion, while Müller et al. (2019) identify parent–child HL communication as a mediator of self-esteem and adaptive psychosocial functioning. Extending this literature, the present results indicate that PTSD resilience emerges from the interactional ecology of home and host-language contexts rather than from discrete linguistic competencies, highlighting the necessity of integrated language environments for trauma mitigation.

On the other hand, CPTSD functional impairment revealed a pronounced three-way interaction among HL proficiency, parental/home-language use, and host-language engagement, B = 6.09, p = .012, 95% CI [1.32, 10.85], delineating a high-risk linguistic configuration. Figure 5 illustrates that non–Nigerian-language speakers experienced escalating CPTSD impairment with increasing HL proficiency (≈2.4 → 5.0), whereas Nigerian-language speakers maintained relatively stable impairment across proficiency levels (≈3.3 → 3.9). Critically, children with high HL proficiency but low host-language engagement and minimal parental/home-language support exhibited elevated CPTSD functional impairment, indicating that HL proficiency in isolation is insufficient as a protective factor.

These patterns corroborate prior work linking HL competence to heritage identity and psychosocial outcomes (Arrendondo et al., 2016; Shin, 2016), but elucidate an ecological boundary condition: the protective potential of HL engagement is contingent upon concurrent host-language engagement and familial support (Lam & Catto, 2023). Consistent with Aksaç (2025), deficits in host-language proficiency exacerbate vulnerability, while cumulative linguistic deprivation predicts maximal CPTSD-related functional impairment. Thus, trauma-related vulnerability in CPTSD is not merely additive but emerges from nested, configuration-dependent linguistic environments.

Indeed, PTSD functional impairment appears primarily modulated by two-way home × host-language interactions, with modest effect magnitude (partial η² = .035). In contrast, CPTSD functional impairment is disproportionately sensitive to three-way HL × parental/home × host-language interactions, reflecting a more complex, configuration-dependent risk landscape (B = 6.09, 95% CI [1.32, 10.85]). Figures 5 and 6 exemplify these dynamics: PTSD resilience requires bi-level linguistic synergy, whereas CPTSD vulnerability is amplified under cumulative deprivation across multiple language layers.

Collectively, these findings advance a nuanced, ecologically grounded understanding of trauma-related functional outcomes, demonstrating that nested language environments—rather than isolated language competencies—shape children’s adaptive trajectories following trauma exposure. Intervention frameworks should therefore prioritize multilayered linguistic scaffolding, promoting both heritage- and host-language engagement. Particular emphasis is warranted for children exhibiting the high-risk CPTSD linguistic profile, wherein deficits across home and host-language domains converge to produce the greatest functional impairment.

Hypothesis 5 examined whether emotional and sexual trauma exposure would predict variation in PTSD and CPTSD functional impairment among refugee children. Multivariate analyses revealed statistically significant effects of emotional trauma, sexual trauma, and their interaction on combined functional impairment outcomes, although effect sizes were modest (partial η² = .019–.028), indicating small but reliable trauma configuration effects. Crucially, univariate analyses demonstrated diagnosis-specific differentiation: CPTSD functional impairment was significantly shaped by emotional trauma and the emotional × sexual trauma interaction, whereas PTSD functional impairment remained comparatively stable across exposure levels. This divergence provides empirical support for the functional distinctiveness of CPTSD in displaced youth populations.

The findings align with the ICD-11 conceptualization of CPTSD as encompassing disturbances in self-organization (DSO), including affect dysregulation, negative self-concept, and relational disturbance (World Health Organization, 2018; Cloitre et al., 2013). These domains extend beyond fear-based PTSD symptom clusters and are theoretically more susceptible to cumulative and interacting forms of trauma. In refugee contexts, children are rarely exposed to single-event trauma; rather, emotional neglect, interpersonal violations, displacement stress, and chronic insecurity often co-occur. The present results suggest that CPTSD-related impairment may operate as a configuration-sensitive construct, responsive to interacting trauma exposures rather than isolated events.

The emotional × sexual trauma interaction is particularly consistent with polyvictimization frameworks, which argue that layered trauma exposures produce multiplicative disruptions in regulatory systems (Finkelhor et al., 2007).

Slope patterns further revealed a nonlinear trajectory for CPTSD functional impairment across emotional trauma levels. Impairment decreased at low exposure levels before increasing at higher levels. This pattern challenges linear dose–response assumptions common in PTSD research and suggests threshold-based adaptation processes. Theoretically, moderate stress exposure can facilitate short-term adaptive recalibration or stress inoculation (Rutter, 2012), whereas higher cumulative exposure may overwhelm affect-regulatory systems, producing disturbances characteristic of CPTSD. In protracted displacement contexts, children may initially mobilize adaptive coping in response to limited adversity; however, escalating emotional trauma likely destabilizes identity and relational systems central to DSO pathology.

Although sexual trauma is strongly associated with PTSD symptomatology in children (e.g., Habigzang et al., 2010), PTSD functional impairment in the present sample did not significantly vary across trauma levels. This divergence underscores an important distinction between symptom presence and functional disruption. In refugee environments characterized by chronic socioeconomic constraints, functional roles may become stabilized by environmental necessity, limiting variance in observable impairment even when symptom severity fluctuates. Thus, PTSD may capture acute fear-based reactivity, whereas CPTSD may better index enduring dysregulation affecting relational and self-structural domains.

Children living in displacement contexts often experience chronic insecurity, disrupted educational trajectories, and social marginalization in addition to direct trauma exposure. Research on displaced populations emphasizes the compounding impact of pre-migration trauma and post-migration stressors (Miller & Rasmussen, 2010). Within such environments, trauma configuration may influence psychosocial functioning more strongly than isolated trauma types. The present findings therefore highlight the importance of assessing cumulative and interacting exposures when evaluating diagnostic outcomes in refugee children.

Collectively, the results suggest several important conceptual advances. CPTSD appears to exhibit greater functional sensitivity to interacting trauma types than PTSD in refugee youth, highlighting its configuration-dependent vulnerability. Trauma-related functional impairment operates conditionally and cumulatively rather than following a simple linear pattern, and functional adaptation trajectories in displaced children may be nonlinear, reflecting both short-term adaptive recalibration and threshold collapse under higher cumulative exposure. Although effect sizes were modest, the consistent interaction and diagnosis-specific patterns indicate that CPTSD represents a structurally differentiated construct with particular relevance for trauma-exposed refugee populations. These findings underscore the value of assessing functional impairment—not solely symptom counts—as a more ecologically valid indicator of trauma-related disruption in displacement contexts.

Hypothesis 6 proposed that trauma exposure, specifically witnessed and physical trauma, would predict CPTSD- and PTSD-related functional impairment among refugee children. Multivariate analyses indicated that witnessed trauma was the primary contributor to combined functional impairment outcomes (Pillai’s Trace = 5.747, F(12, 610) = 5.747, p < .001, partial η² = .102), whereas physical trauma exerted a smaller, yet statistically significant effect (F(10, 610) = 1.881, p = .045, partial η² = .030). Univariate analyses further delineated these effects: witnessed trauma predicted CPTSD functional impairment with a large effect (F(6, 305) = 9.201, p < .001, partial η² = .153) and PTSD impairment with a moderate effect (F(6, 305) = 4.160, p = .042, partial η² = .076), whereas physical trauma demonstrated modest or trend-level associations. Examination of estimated marginal means (Figures 9 and 10) revealed that CPTSD impairment increased sharply from no exposure to initial witnessed trauma (Δ ≈ 2 points), peaked at moderate exposure levels (estimated means 3.7–4.6), and stabilized thereafter. Physical trauma contributed additive effects across all levels of witnessed trauma without altering the slope, suggesting that its impact is consistent but secondary to witnessed trauma.

**Figure 10.**
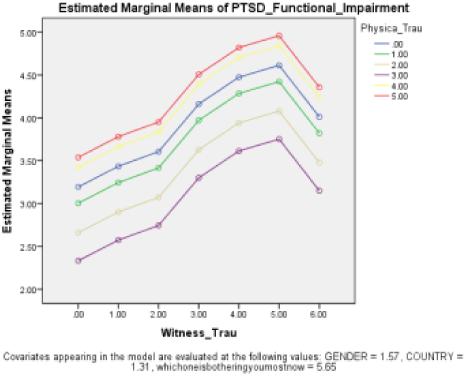
Estimated Marginal Means of PTSD Functional Impairment by Witnessed Trauma and Physical Trauma.

These findings align with extant literature indicating that trauma exposure—particularly vicarious or witnessed trauma—exerts profound biopsychosocial effects in children and adolescents. PTSD in youth disrupts emotional regulation, behavioral control, social engagement, and academic functioning, increasing the risk for long-term psychosocial maladjustment (Russo et al., 2023; Motsan, Yirmiya, & Feldman, 2022). Witnessed trauma, often chronic and relationally embedded, may disproportionately compromise affective regulation and interpersonal functioning, which are central to CPTSD symptomatology, thereby corroborating the ICD-11 distinction between PTSD and CPTSD (Cloitre et al., 2018; Karatzias et al., 2019).

The plateau observed at higher levels of witnessed trauma reflects threshold and stress saturation effects: initial exposure appears sufficient to trigger substantial functional disruption, while subsequent exposures maintain high impairment without proportional escalation. This pattern is consistent with allostatic load models, which posit that repeated environmental stressors dysregulate neuroendocrine and immune systems, producing sustained functional deficits (Warren, 2025; Hassanpour, 2023). Physiological sequelae—including sleep disruption, somatic complaints, impaired immune functioning, headaches, gastrointestinal disturbances, and growth retardation—documented in trauma-exposed children further support this biopsychosocial interpretation (Rahman, Mensah, & Oladele, 2025).

The differential effects of trauma type were notable. While physical trauma exerted statistically significant but smaller additive effects, it did not modify the trajectory of witnessed trauma-related impairment. This pattern suggests that indirect or observational trauma may exert more pervasive developmental consequences than direct physical harm, particularly in refugee contexts characterized by chronic environmental threat. The near-parallel trajectories of impairment across physical trauma levels indicate additive rather than interactive effects, underscoring the dominant role of witnessed trauma in shaping functional outcomes.

Collectively, these findings highlight the centrality of witnessed trauma in driving functional impairment among refugee children, with CPTSD showing heightened sensitivity compared to PTSD. They refine prior models by elucidating diagnosis-specific vulnerability, additive versus interactive trauma effects, and exposure-threshold patterns. Importantly, these results emphasize the critical need for early, trauma-informed interventions that target both emotional regulation and relational functioning, aiming to mitigate long-term psychosocial and developmental consequences in pediatric refugee populations.

### Implications for Theory, Policy, and Practice

Grounded in ethnolinguistic theory and ecosystem (ecological systems) theory, this study demonstrates that trauma-related functional impairment in children emerges from differentiated interactions between cumulative trauma exposure and contextual resources embedded within family, educational, and sociocultural environments. From an ecosystem perspective, PTSD and CPTSD represent distinct functional pathways shaped by disruptions at different ecological levels. PTSD-related impairment appears more responsive to proximal microsystem and mesosystem resources—particularly family language practices and school-based linguistic inclusion—whereas CPTSD reflects deeper, cumulative disruptions associated with chronic exposure to violence that overwhelm such contextual supports.

Ethnolinguistic theory clarifies why language-related factors function as conditional protective mechanisms rather than universal buffers. Heritage-language continuity and meaningful linguistic engagement within the family serve as relational and identity-stabilizing resources that may support emotional regulation and functional adaptation in children whose trauma exposure is limited or episodic. In contrast, when trauma is cumulative and interpersonal—as reflected in CPTSD—these resources appear insufficient to offset impairment, underscoring the primacy of trauma severity and chronicity over cultural or linguistic context.

The demographic patterns observed in this study further illuminate these processes. Widespread erosion of functional heritage-language competence, coupled with partial retention of affective attachment to parental languages, suggests that many children inhabit linguistically fragmented environments. Progressive shifts toward host-environment languages across age groups indicate intensifying linguistic assimilation with prolonged displacement, potentially compounding identity disruption and constraining the long-term protective value of heritage-language resources. Variation in home-language use across religious groups highlights the socially patterned nature of linguistic environments, reflecting differences in community integration and language maintenance practices rather than direct psychological effects.

### Alignment with SDGs, Child Protection, and Humanitarian Programming

These findings contribute directly to SDG 3 (Good Health and Well-Being) by identifying differentiated mechanisms of trauma-related functional impairment, supporting the development of trauma-informed mental health and psychosocial support (MHPSS) interventions that are responsive to trauma severity and exposure profiles. The selective relevance of language-related factors for PTSD—but not CPTSD—reinforces the need for stratified, diagnosis-sensitive approaches to child mental health in displacement settings.

The study also advances SDG 4 (Quality Education) by demonstrating that linguistically inclusive and culturally responsive educational environments can function as protective mesosystem conditions for children with PTSD-related impairment. Schools emerge as critical ecological settings where language practices may either buffer or exacerbate functional difficulties, depending on the child’s trauma profile.

In relation to SDG 16 (Peace, Justice, and Strong Institutions), the strong association between cumulative trauma exposure and CPTSD-related functional impairment highlights the long-term developmental consequences of violence, instability, and weak protective systems. These findings underscore that exposure to violence produces enduring functional harm that cannot be mitigated solely through microsystem-level supports such as family language continuity, reinforcing the importance of structural prevention and protection mechanisms.

Consistent with child protection frameworks, the results demonstrate that CPTSD-related impairment reflects cumulative ecological harm rather than isolated adverse events. While cultural and linguistic resources may enhance resilience in some children, they cannot substitute for sustained safety, relational repair, and trauma-specific care in cases of chronic exposure. This supports child protection priorities emphasizing early identification, case management, and restoration of safe caregiving environments.

For humanitarian programming, the findings align with layered MHPSS models. Children with PTSD-related impairment may benefit from community- and school-based interventions that strengthen family engagement and language-inclusive practices, whereas children with CPTSD require higher-tier, specialized services that address relational disruption and chronic trauma exposure. These results caution against one-size-fits-all psychosocial interventions and support ecologically stratified service delivery based on trauma profiles.

### Limitations and Future Directions

Several limitations should be noted. First, the cross-sectional design precludes causal inference regarding the developmental sequencing of language erosion, cumulative trauma exposure, and functional impairment. Longitudinal research is needed to examine how linguistic environments and trauma exposure interact dynamically across developmental stages. Second, reliance on self-and caregiver-reported measures may introduce recall bias. Third, the focus on Liberian and Sierra Leonean refugee children in Nigeria may limit generalizability to other displacement contexts.

Unmeasured cognitive and environmental variables, including caregiver mental health, educational quality, and executive functioning, likely interact with both language and trauma-related outcomes and warrant further investigation. Finally, while multivariate analyses revealed robust main effects, some higher-order interaction effects were modest in magnitude. Importantly, these small effect sizes reinforce rather than undermine the theoretical conclusion that CPTSD-related impairment is driven primarily by cumulative trauma exposure rather than contextual modifiers such as language.

## Conclusion

This study provides compelling evidence that PTSD and CPTSD are associated with distinct patterns of functional impairment shaped by different ecological and experiential determinants. Language-related characteristics and family language practices play a meaningful role in shaping functional outcomes in PTSD, consistent with theories emphasizing ecological and cultural resources. In contrast, CPTSD-related impairment is predominantly determined by cumulative trauma exposure—particularly witnessing and physical trauma—supporting conceptualizations of CPTSD as a condition with qualitatively different developmental pathways rather than a more severe variant of PTSD.

By integrating ethnolinguistic and ecosystem perspectives, the study advances trauma theory beyond symptom-based models and demonstrates the necessity of developmentally, diagnostically, and ecologically informed approaches to research, intervention, and policy. Effective responses to trauma-related functional impairment in children must therefore balance cultural and linguistic sensitivity with sustained attention to the severity, chronicity, and relational nature of traumatic exposure.

### Overall Significance

Scientifically, the findings strengthen the empirical distinction between PTSD and CPTSD by linking diagnostic differentiation to functional and ecological mechanisms. Methodologically, the study illustrates the value of multivariate approaches for disentangling overlapping trauma responses while accounting for contextual moderators. Practically and at the policy level, the results support integrated child protection and MHPSS strategies that combine language-inclusive education with trauma-specific mental health care, particularly for children exposed to chronic violence.

Taken together, this study offers a theoretically grounded, context-sensitive contribution to the understanding of childhood trauma in displacement settings and provides a robust evidence base for differentiated, ecosystem-informed interventions.

## Data Availability Statement

The datasets generated and analyzed during the current study are publicly available in the **Zenodo repository** at 10.5281/zenodo.15817238.

## Data Availability

All data produced in this present study are available online at: 10.5281/zenodo.15817238

10.5281/zenodo.15817238

## References

Aguilar, M., Ferré, P., & Hinojosa, J. A. (2024). The landscape of emotional language processing in bilinguals: A review. Psychology of Learning and Motivation, 80, 1–32. 10.1016/bs.plm.20

Aksaç, T. (2025). Experiences of Syrian refugee children learning Turkish as a foreign language at the primary school level. The Primary Education Journal. Retrieved from https://primedu.org

American Psychiatric Association. (2013). Diagnostic and statistical manual of mental disorders (5th ed.). American Psychiatric Publishing.

Anderson, A., Hamilton, R., Moore, D., Loewen, S., & Frater-Mathieson, K. (2004). Education of refugee children: Theoretical perspectives and best practice. In R. Hamilton & D. Moore (Eds.), Educational interventions for refugee children (pp. 15–25). Routledge.

Arrendondo, M. M., Rosado, M., & Satterfield, T. (2016). Understanding the impact of heritage language on ethnic identity formation and literacy for U.S. Latino children. Journal of Cognition and Culture, 16, 245–266. 10.1163/15685373-12342179

Badecker, W., & Kuminiak, F. (2007). Morphology, agreement and working memory retrieval in sentence production: Evidence from gender and case in Slovak. Journal of Memory and Language, 56(1), 65–85. 10.1016/j.jml.2006.08.004

Bailen, N. H., Green, L. M., & Thompson, R. J. (2019). Understanding emotion in adolescents: A review of emotional frequency, intensity, instability, and clarity. Emotion Review, 11(1), 63–73. 10.1177/1754073918768878

Baker, M. (1993). Corpus: Linguistics and Translation Studies: Implications and Applications. In M. Baker, G. Francis, & E. Tognini-Bonelli (Eds.), Text and Technology: In honor of John Sinclair. Amsterdam (pp. 233–250). John Benjamins. 10.1075/z.64.15bak

Bartels, L., Solheim Skar, A.-M., Skogbrott Birkeland, M., Ormhaug, S. M., Berliner, L., & Jensen, T. K. (2023). The differential impact of DSM-5 post-traumatic stress symptoms on functional impairment in traumatized children and adolescents. European Child & Adolescent Psychiatry, 33(5), 1573–1581. 10.1007/s00787-023-02266-w (pmc.ncbi.nlm.nih.gov)

Becker, A. & Magno, C. (2022). Cognitive migration through language: Capturing linguistic movement and barriers in language portraits. In C. Magno, J. Lew, & S. Rodriguez (Eds.), (Re)Mapping migration and education: Centering methods and methodologies (pp. 134–157). Brill.

Betancourt TS, Borisova I, Williams TP, Meyers-Ohki SE, Rubin-Smith JE, Annan J, Kohrt BA. Psychosocial adjustment and mental health in former child soldiers: a systematic review of the literature and recommendations for future research. J Child Psychol Psychiatry

Boye, K., Bastiaanse, R., Harder, P., & Martínez-Ferreiro, S. (2023). Agrammatism in a usage-based theory of grammatical status: Impaired combinatorics, compensatory prioritization, or both? Journal of Neurolinguistics, 65, 101108. 10.1016/j.jneuroling.2022.101108

Bronfenbrenner, U. (1977). Toward an experimental ecology of human development. American Psychologist, 32(7), 513–531. 10.1037/0003-066X.32.7.513

Bronfenbrenner, U. (1979). The ecology of human development: Experiments by nature and design. Harvard University Press.

Bronfenbrenner, U., & Morris, P. A. (1998). The bioecological model of human development. In R. M. Lerner (Ed.), Handbook of child psychology (pp. 993–1027). Wiley & Sons.

Bronfenbrenner, U., & Morris, P. A. (2006). The bioecological model of human development. In R. M. Lerner (Ed.), Handbook of child psychology (6th ed., Vol. 1, pp. 793–828). John Wiley & Sons, Inc. 10.1002/9780470147658.chpsy0114

Brown, C. L. (2009). Heritage language and ethnic identity: A case study of Korean-American college students. International Journal of Multicultural Education, 11(1), 1–16. 10.18251/ijme.v11i1.157

Brumberg, Stephen F. 1986. Going to America, Going to School: The Jewish Immigrant Public School Encounter in Turn-of- The-Century New York City. New York: Praeger.

Caldwell-Harris, C. L. (2014). Emotionality differences between a native and foreign language: Theoretical implications. Frontiers in Psychology, 5, 1055. 10.3389/fpsyg.2

Chen, X., & Padilla, A. M. (2019). Role of bilingualism and biculturalism as assets in positive psychology: Conceptual dynamic GEAR model. Frontiers in Psychology, 10, 2019. 10.3389/fpsyg.2019.02122

Chiswick, B. R., & Miller, P. W. (2002). Immigrant earnings: Language skills, linguistic concentrations and the business cycle. Journal of Population Economics, 15(1), 31–57. 10.1007/PL00003838

Ciolino, C., Hyter, Y. D., Suarez, M., & Bedrosian, J. (2021). Narrative and other pragmatic language abilities of children with a history of maltreatment. Perspectives of the ASHA Special Interest Groups, 6(2), 230–241. 10.1044/2020_PERSP-20-00136

Cohodes, E.M., Kribakaran, S., Odriozola, P., Bakirci, S., McCauley, S., Hodges, H.R., Sisk, L.M., Zacharek, S.J., & Gee, D.G. (2021). Migration-related trauma and mental health among migrant children emigrating from Mexico and Central America to the United States: Effects on developmental neurobiology and implications for policy. Developmental Psychobiology, 63(6), e22158. 10.1002/dev.22158

Costa, A., et al. (2021). The role of emotional resonance in multilingual emotional expression. Journal of Psycholinguistic Research, 50(5), 825–840

Cross, D., Fani, N., Powers, A., & Bradley, B. (2017). Neurobiological development in the context of childhood trauma. Clinical Psychology: Science and Practice, 24(2), 111–124. 10.1111/cpsp.12198

Cummins, Jim. (1978). “Metalinguistic Development of Children in Bilingual Education Programs.”

Cutler, S. A., & Dwyer, D. J. (1981). Maniyaka: A reference handbook of phonetics, grammar, lexicon and learning procedures (ED247767). ERIC Clearinghouse. https://files.eric.ed.gov/fulltext/ED247767.pdf

DeCamp D. Pidgins and creoles as standard languages. In: Baker P, Winer L, editors. From Contact to Creole and Beyond: Language and Change in Africa and the Caribbean. Kingston: University of the West Indies Press; 2012. p. 45–63.

Deldar, Z., Gevers-Montoro, C., Khatibi, A., & Ghazi-Saidi, L. (2020). The interaction between language and working memory: A systematic review of fMRI studies in the past two decades. AIMS Neuroscience, 8(1), 1. 10.3934/Neuroscience.2021001

Dunn, E. C., Nishimi, K., Powers, A., & Bradley, B. (2017). Is developmental timing of trauma exposure associated with depressive and post-traumatic stress disorder symptoms in adulthood? Journal of Psychiatric Research, 84, 119–127. 10.1016/j.jpsychires.2016.10.015 (PMCID: PMC5479490)

Durodola TS. Narratives of the journey to exile and transformative agency of residual Liberian refugees in Oru, southwestern Nigeria [master’s thesis]. Ibadan (NG): Institute of African Studies, University of Ibadan; 2021.

Dye, H. (2018). The impact and long-term effects of childhood trauma. Journal of Human Behavior in the Social Environment, 28(3), 381–392. 10.1080/10911359.2018.1435328

Fan, L., & Kang, T. (2025). Early childhood trauma and its long-term impact on cognitive and emotional development: A systematic review and meta-analysis. Annals of Medicine, 57(1), 2536199. 10.1080/07853890.2025.2536199

Fan, L., & Kang, T. (2025). Early childhood trauma and its long-term impact on cognitive and emotional development: A systematic review and meta-analysis. Annals of Medicine, 57(1), Article 2536199. 10.1080/07853890.2025.2536199

Fatima, N., Afzaal, H. M., Hussain, Z., & Sajid, M. K. M. (2024). Language and emotion: A study of emotional expression in multilinguals. Journal of Applied Linguistics and TESOL, 7(4).

Fedorenko, E., Ryskin, R., & Gibson, E. (2023). Agrammatic output in non-fluent, including Broca’s, aphasia as a rational behavior. Aphasiology, 37(12), 1981–2000. 10.1080/02687038.2022.2143233

Ferré, P., Fraga, I., & Hinojosa, J. A. (2025). The interplay between language and emotion: Introduction to the special issue. Cognition and Emotion, 39(7), 1405–1417. 10.1080/02699931.2025.2549966

Ferré, P., Fraga, I., & Hinojosa, J. A. (2025). The interplay between language and emotion: A narrative review. Cognition and Emotion.

Fields-Olivieri, M. A., Kim, Y., Jennings, K. J., & Cole, P. M. (2024). The role of language in the development of emotion regulation. In M. A. Bell (Ed.), Child development at the intersection of emotion and cognition (2nd ed.) (pp. 31–50). American Psychological Association. 10.1037/0000406-003

Filippi, R., Ceccolini, A., Booth, E., Shen, C., Thomas, M., Toledano, M., & Dumontheil, I. (2022). Modulatory effects of SES and multilinguistic experience on cognitive development: A longitudinal data analysis of multilingual and monolingual adolescents from the SCAMP cohort. International Journal of Bilingual Education and Bilingualism, 25(9), 3489–3506. 10.1080/13670050.2022.2064191

Finkelhor, D., Ormrod, R. K., & Turner, H. A. (2007). Poly-victimization: a neglected component in child victimization. Child Abuse & Neglect, 31(1), 7–26. 10.1016/j.chiabu.2006.06.008

Fishman, J. (1966) 1966. Hungarian language maintenance in the United States. Bloomington: Indiana University Press

Ford, P.R., Yates, I., & Williams, A.M. (2010). An analysis of practice activities and instructional behaviours used by youth soccer coaches during practice: Exploring the link between science and application. Journal of Sports Sciences, 28(5), 483–495. 10.1080/02640410903582750

Fornell, C., & Larcker, D. F. (1981). Structural Equation Models with Unobservable Variables and Measurement Error: Algebra and Statistics. Journal of Marketing Research, 18, 382–388. 10.2307/3150980

Fridrikh, A., Pentón Herrera, L. J., & Kałdonek-Crnjaković, A. (2025). “English was a real help”: Multilingualism as an integration strategy among Ukrainian refugee children in Poland. International Journal of Bilingualism. Advance online publication. 10.1177/13670069251362740

Fyle, C. M. (1994). Official and unofficial attitudes and policy towards Krio as the main lingua franca in Sierra Leone. In R. Fardon & G. Furniss (Eds.), African languages, development, and the state (pp. 44–54). Routledge.

Giles, H., & Johnson, P. (2010). Ethnolinguistic identity theory. In R.L. Jackson II & M.A. Hogg (Eds.), Encyclopedia of identity (Vol.111). Sage Publications.

Gillam, S. L., & Hyter, Y. (2025). Developing trauma-informed competence in speech-language pathologists: Foundations for narrative assessment and intervention. Perspectives of the ASHA Special Interest Groups, 10, 1–11. 10.1044/2025_PERSP-25-00073

Gillam, S., Gillam, R., Magimairaj, B., Capin, P., IsraelsenAugenstein, M, Roberts, G., & Vaughn, S. (2024). Contextualized, multi-component language instruction: From theory to randomized controlled trial. Language, Speech and Hearing Services in Schools, 55(3), 661–682. 10.1044/2024_LSHSS-23-00171

Graham-Bermann, S. A., Gruber, G., Howell, K. H., & Girz, L. (2009). Factors discriminating among profiles of resilience and psychopathology in children exposed to intimate partner violence (IPV). Child Abuse & Neglect, 33(9), 648–660. 10.1016/j.chiabu.2009.01.002

Grasso, D. J., Ford, J. D., & Briggs-Gowan, M. J. (2012). Early life trauma exposure and stress sensitivity in young children. Journal of Pediatric Psychology, 38(1), 94–103. 10.1093/jpepsy/jss101 (PMC)

Habigzang, L. F., Borges, J. L., Dell’Aglio, D. D., & Koller, S. H. (2010). Characterization of posttraumatic stress disorder (PTSD) symptoms in sexually abused girls. Psicologia: Reflexão e Crítica, 22(1), 27–44. 10.1590/S0102-79722009000100004

Hakuta, K., & Diaz, R. M. (1985). The relationship between degree of bilingualism and cognitive ability: A critical discussion and some new longitudinal data. In K. E. Nelson (Ed.), Children’s language, Vol. 5, pp. 319–344). Lawrence Erlbaum Associates

Hakuta, Kenji.(1985). “The Relationship Between Bilingualism and Cognitive Ability: A Critical Discussion and Some Longitudinal Data.”

Hamuddin, B., Ramadhani, M. R., & Ningrum, F. S. (2025). The impact of home language and literacy environment on children’s learning outcomes. Script Journal: Journal of Linguistics and English Teaching, 10(1), 138–163. 10.24903/sj.v10i1.2001

Hansegård, N. E. (1968). Tvåspråkighet eller halvspråkighet? Aldus/Bonniers.

Haselgruber, A., Sölva, K., & Lueger-Schuster, B. (2020a). Validation of ICD-11 PTSD and Complex PTSD in foster children using the International Trauma Questionnaire. Acta Psychiatrica Scandinavica, 141(1), 60–73. 10.1111/acps.13100

**Hassanpour**, **H**. (2023). Interaction of allostatic load with immune, inflammatory, and coagulation systems. Immunoregulation, 5(2), 91–100. 10.32598/Immunoregulation.5.2.4

Herd, T., King-Casas, B., & Kim-Spoon, J. (2020). Developmental changes in emotion regulation during adolescence: Associations with socioeconomic risk and family emotional context. Journal of Youth and Adolescence.

Ho, G. W. K., Liu, H., Karatzias, T., Hyland, P., Cloitre, M., Lueger-Schuster, B., Brewin, C. R., Guo, C., Wang, X., & Shevlin, M. (2022). Validation of the International Trauma Questionnaire-Child and Adolescent Version (ITQ-CA) in a Chinese mental health service seeking adolescent sample. Child and Adolescent Psychiatry and Mental Health, 16(1), 66. 10.1186/s13034-022-00497-4

Hopfinger, L., Berking, M., Bockting, C. L. H., et al. (2016). Emotion regulation mediates the effect of childhood trauma on depression. Journal of Affective Disorders, 198, 189–197. 10.1016/j.jad.2016.03.050

Horn, R., Arakelyan, S., Wurie, H., & Ager, A. (2021). Factors contributing to emotional distress in Sierra Leone: a socio-ecological analysis. International Journal of Mental Health Systems, 15(1), Article 58. 10.1186/s13033-021-00474-y

Huberty, C. J., & Morris, J. D. (1989). Multivariate analysis versus multiple univariate analyses. Psychological Bulletin, 105(2), 302–308. 10.1037/0033-2909.105.2.302

Hyter, Y. D. (2021). Childhood maltreatment consequences on social pragmatic communication: A systematic review of the literature. Perspectives of the ASHA Special Interest Groups, 6(2), 262–287. 10.1044/2021_PERSP-20-00222

Isen, L. E., & Boye, K. (2025). Language in PTSD: Evidence for linguistic impairment in two case studies. Studies in Language and Mind, [volume]([issue]), [page range]. 10.19090/slm.6.7

Kang, H. S. (2013). Korean American college students’ language practices and identity positioning: ‘Not Korean, but not American’. Journal of Language, Identity, and Education, 12(4), 248– 261. 10.1080/15348458.2013.818473

Karlander, D., & Salö, L. (2026). Semilingualism — a cringe concept in bilingualism studies and beyond. ResearchGate. https://www.researchgate.net/publication/399664000_Semilingualism_-_a_cringe_concept_in_bilingualism_studies_and_beyond

Kazlauskas, E., Zelviene, P., Daniunaite, I., Hyland, P., Kvedaraite, M., Shevlin, M., & Cloitre, M. (2020). The structure of ICD-11 PTSD and Complex PTSD in adolescents exposed to potentially traumatic experiences. Journal of Affective Disorders, 265, 169–174. 10.1016/j.jad.2020.01.061

Kolk, H. (1995). A time-based approach to agrammatic production. Brain and Language, 50(3), 282– 303. 10.1006/brln.1995.1049

Kunz E. Exile and resettlement: Refugee theory. Int Migr Rev. 1981;15(1):42–51

Lam, V. L., & Catto, A. C. (2023). Heritage Language Use and Proficiency: Acculturation, Identities and Psychological Health. Journal of Home Language Research, 6(1): 3, pp.1–19. DOI: 10.16993/jhlr.51

Le, M. T. H., Holton, S., Romero, L., & Fisher, J. (2018). Polyvictimization among children and adolescents in low- and lower-middle-income countries: A systematic review and meta-analysis. *Trauma, Violence*, & Abuse, 19(3), 323–342. 10.1177/1524838016662707

Leclerc J. Aménagement linguistique en Afrique: Libéria [Internet]. 2002 [cited 2025 Sep 22]. Available from: https://www.axl.cefan.ulaval.ca/afrique/liberia.htm

Leopold, W. F. (1961). Patterning in children’s language learning. In Saporta (ed.), (1961). 350–358.Google Scholar

Lewis, S. J., Arseneault, L., Caspi, A., Fisher, H. L., Matthews, T., Moffitt, T. E., & Danese, A. (2019). The epidemiology of trauma and post-traumatic stress disorder in a representative cohort of young people in England and Wales. The Lancet Psychiatry, 6(3), 247–256. 10.1016/S2215-0366(19)30031-8

Lindner, K., Hipfner-Boucher, K., Yamashita, A., Riehl, C. M., Ait Ramdan, M., & Chen, X. (2020). Acculturation through the lens of language: Syrian refugees in Canada and Germany. Applied Psycholinguistics, 41(6), 1351–1374. 10.1017/S0142716420000454

Liu, L. L, Benner, A. D., Lau, A. S., & Kim, S. Y. (2009). Mother-adolescent language proficiency and adolescent academic and emotional adjustment among Chinese American families. Journal of Youth and Adolescence, 38(4), 572–586. 10.1007/s10964-008-9358-8

Llanes, A., et al. (2020). The impact of multilingualism on emotional expression and regulation. Bilingualism: Language and Cognition, 23(2), 207–218.

Lupindo, M.B., Lorenz, H., French, S., & Salkovskis, P. (2025). Impact of exposure to community and school violence during adolescence in the African context: Systematic review. The British Journal of Psychiatry International. 10.1192/bji.2025.10043

Maciejewski DF, van Lier PAC, Branje SJT, Meeus WHJ, & Koot HM (2015). A 5-year longitudinal study on mood variability across adolescence using daily diaries. Child Development, 86, 1908–1921. doi: 10.1111/cdev.12420 [DOI] [PubMed] [Google Scholar][Ref list]

Majid, A. (2012). Current emotion research in the language sciences. Emotion Review, 4(4), 432–443. 10.1177/1754073912445827

Malarbi S., Abu-Rayya H. M., Muscara F., Stargatt R. (2017). Neuropsychological functioning of childhood trauma and post-traumatic stress disorder: A meta-analysis. Neuroscience and Biobehavioral Reviews, 72, 68–86. 10.1016/j.neubiorev.2016.11.004 [DOI] [PubMed] [Google Scholar

Martinovic, Borja, Frank van Tubergen and Ineke Maas. 2009. “Dynamics of Interethnic Contact: A Panel Study of Immigrants in the Netherlands.” European Sociological Review 25(3):303–18.

McKinley, A. C. (2026). Theoretical insights into cumulative childhood adversity and homicide victimization: A conditional vulnerability approach. In I. Bryce & S. Collier (Eds.), Cumulative childhood harm in family law and criminal justice systems (pp. 187–216). IGI Global. 10.4018/979-8-3373-9524-1.ch007

Miller, K. E., & Rasmussen, A. (2010). War exposure, daily stressors, and mental health in conflict and post-conflict settings: Bridging the divide between trauma-focused and psychosocial frameworks. Social Science & Medicine, 70(1), 7–16. 10.1016/j.socscimed.2009.09.029

Motsan, S., Yirmiya, K., & Feldman, R. (2022). Chronic early trauma impairs emotion recognition and executive functions in youth: Specifying biobehavioral precursors of risk and resilience. Development and Psychopathology, 34, 1339–1352. 10.1017/S0954579421000067

Motti-Stefanidi, F., & Masten, A. S. (2017). A resilience perspective on immigrant youth adaptation and development. In F. Motti-Stefanidi & A. S. Masten (Eds.), Handbook on positive development of minority children and youth (pp. 19–34). Springer International Publishing. 10.1007/978-3-319-43645-6_2

Müller, L. M., Howard, K., Wilson, E., Gibson, J., & Katsos, N. (2019). Bilingualism in the family and child well-being: A scoping review. International Journal of Bilingualism, 24(5–6), 1049–1070. 10.1177/1367006920920939

Murphy, F., Nasa, A., Cullinane, D., Raajakesary, K., Gazzaz, A., Sooknarine, V., Haines, M., Roman, E., Kelly, L., O’Neill, A., Cannon, M., & Roddy, D. W. (2022). Childhood trauma, the HPA axis and psychiatric illnesses: A targeted literature synthesis. Frontiers in Psychiatry, 13. 10.3389/fpsyt.2022.748372

Naughton, C., Drennan, J., Hyde, A., Allen, D., O’Boyle, K., Felle, P., & Butler, M. (2013). An evaluation of the appropriateness and safety of nurse and midwife prescribing in Ireland. Journal of Advanced Nursing, 69(7), 1478–1488. 10.1111/jan.12004

Nelson, C. A., Scott, R. D., Bhutta, Z. A., Burke Harris, N., Danese, A., & Samara, M. (2020). Adversity in childhood is linked to mental and physical health throughout life. BMJ, 371, Article m3048. 10.1136/bmj.m3048

Niles, A. N., Craske, M. G., Lieberman, M. D., & Hur, C. (2015). Affect labeling enhances exposure effectiveness for public speaking anxiety. Behaviour Research and Therapy, 68, 27– 36. 10.1016/j.brat.2015.03.004

Nook, E. C., Sasse, S. F., Lambert, H. K., McLaughlin, K. A., & Somerville, L. H. (2017). Increasing verbal knowledge mediates development of multidimensional emotion representations. Nature Human Behaviour, 1(12), 881–889. 10.10

Norte, C. E., Vargas, A. L. V., & de Carvalho Silveira, A. (2024). Post-traumatic stress disorder and working memory: A systematic review. Trends in Psychology, 32(2), 612–623. 10.1007/s43076-022-00206-2

Nwagbo OG. Identity and code switching among Liberian refugees in Oru Camp, Nigeria. Ihafa: A Journal of African Studies. 2016 Jun;8(1):137–53.

Olson, C. L. (1974). Comparative robustness of six tests in multivariate analysis of variance. Journal of the American Statistical Association, 69(348), 894–908. 10.1080/01621459.1974.10480224

Osunkoya OA. The United Nations and the management of Liberian refugees in West Africa, 1990–2007: A discourse. Wukari Int Stud J [Internet]. 2025 Jun [cited 2025 Sep 20];9(2):191–191. Available from: https://wissjournals.com.ng/index.php/wiss/article/view/629/560

Peal, E., & Lambert, W. E. (1962). The relation of bilingualism to intelligence. Psychological Monographs: General and Applied, 76(27), 1–23. 10.1037/h0093840

Pemberton-Roben, C. K., Bass, A. J., Moore, G. A., Murray-Kolb, L., Tan, P. Z., Gilmore, R. O., Buss, K. A., Cole, P. M., & Teti, L. O. (2012). Let me go: The influences of crawling experience and temperament on the development of anger expression. Infancy, 17(5), 558–577. 10.1111/j.1532-7078.2011.00

Peterson, G. W., & Bush, K. R. (2012). Conceptualizing cultural influences on socialization: Comparing parent–adolescent relationships in the United States and Mexico. In G. W. Peterson & K. R. Bush (Eds.), Handbook of marriage and the family (pp. 177–208). Springer. 10.1007/978-1-4614-3987-5_9

Pollmann, A., Rakesh, D., & Fuhrmann, D. (2025). Longitudinal associations between adolescent adversity, brain development and behavioural and emotional problems. Developmental Cognitive Neuroscience, 77, 101646. 10.1016/j.dcn.2025.101646

Ponari, M., Norbury, C. F., & Vigliocco, G. (2020). The role of emotional valence in learning novel abstract concepts. Developmental Psychology, 56(10), 1855–1865.

Portes A, & Lingxin Hao, 1998. “E Pluribus Unum: Bilingualism and Language Loss in the Second Generation,” Macroeconomics 9805006, University Library of Munich, Germany.

Portes, and Rumbaut.(1996). Immigrant America, A Portrait, 2nd ed. Berkeley, Ca: University of California Press.

Rahman, A., Mensah, D. K., & Oladele, O. K. (2025). Biopsychosocial determinants of health outcomes in life-threatening diseases [Manuscript]. ResearchGate. https://www.researchgate.net/publication/399392385_Biopsychosocial_Determinants_of_Health_Outcomes_in_Life-_Threatening_Diseases

Redican, E., Hyland, P., Cloitre, M., McBride, O., Karatzias, T., Murphy, J., Bunting, L., & Shevlin, M. (2022). Prevalence and predictors of ICD-11 Posttraumatic Stress Disorder and Complex PTSD in young people. Acta Psychiatrica Scandinavica, 146(2), 110–125. 10.1111/acps.13442

Ruba, A. L., Harris, L. T., & Wilbourn, M. P. (2021). Examining preverbal infants’ ability to map labels to facial configurations. Affective Science, 2(2), 142–149. 10.1007/ s42761-

Rumbaut, R, G.(1995). “The New Californians: Comparative Research Findings on the Educational Progress of Immigrant Children.” In R.G. Rumbaut and W.A. Cornelius (eds.) California’s Immigrant Children: Theory, Research, and Implications for Educational Policy. La Jolla, CA.: Center for U.S.-Mexican Studies, University of California-San Diego.

Russo, J. E., Dhruve, D. M., & Oliveros, A. D. (2023). Childhood trauma and PTSD symptoms: Disentangling the roles of emotion regulation and distress tolerance. Research on Child and Adolescent Psychopathology, 51, 1273–1287. 10.1007/s10802-023-01048-x

Sabater, L., Ponari, M., Haro, J., Fernández-Folgueiras, U., Moreno, E. M., Pozo, M. A., Ferré, P., & Hinojosa, J. A. (2023). The acquisition of emotion-laden words from childhood to adolescence. Current Psychology, 42(33), 29280–29290. 10.1007/s12144-022-03989-w

Sachser, C., Berliner, L., Holt, T., Jensen, T. K., Jungbluth, N., Risch, E., … & Goldbeck, L. (2017). International development and psychometric properties of the Child and Adolescent Trauma Screen (CATS). Journal of affective disorders, 210, 189–195. https://pubmed.ncbi.nlm.nih.gov/28049104/

Safiullin, A., & Zheltukhina, M. (2020). Language choice and emotional comfort in multilinguals. Language and Emotion Journal, 12(3), 211–225

Schwieter, J. W., & Sunderman, G. (2020). Emotional expression in second language use: The role of proficiency and familiarity. Language Learning, 70(1), 168–191.

Scontras, G., Fuchs, Z., & Polinsky, M. (2015). Heritage language and linguistic theory. Frontiers in Psychology, 6, 1545. 10.3389/fpsyg.2015.01545

Serrano, A., et al. (2020). The impact of multilingualism on emotional expression. Bilingualism: Language and Cognition, 23(2), 200–216

Sharma, N. (2026). Long term effects of childhood exposure to violence in fragile and conflict affected settings. The BMJ, 392, e086040. 10.1136/bmj-2025-086040

Shelton, L. G. (2019). The Bronfenbrenner primer: A guide to develecology. Routledge. 10.4324/9781315136066

Shields, Michael A., and Stephen Wheatley Price. 2002. “The English Language Fluency and Occupational Success of Ethnic Minority Immigrant Men Living in English Metropolitan Areas.” Journal of Population Economics 15(1):137–60.

Shin, S. J. (2016). Hyphenated identities of Korean heritage language learners: marginalization, colonial discourses and internalized whiteness. Journal of Language, Identity and Education, 15(1), 32–43. 10.1080/15348458.2016.1113815

Shonkoff, J. P., Siegel, B. S., Dobbins, M. I., Earls, M. F., Garner, A. S., McGuinn, L., Pascoe, J., & Wood, D. L. (2012). The lifelong effects of early childhood adversity and toxic stress. Pediatrics, 129(1), e232–e246. 10.1542/peds.2011-2663

Shonkoff, J.P., Slopen, N., & Williams, D.R. (2021). Early childhood adversity, toxic stress, and the impacts of racism on the foundations of health. Annual Review of Public Health, 42(1), 115–134. 10.1146/annurev-publhealth-090419-101940

Silvers, J. A. (2022). Adolescence as a pivotal period for emotion regulation development. Current Opinion in Psychology, 44, 258–263. 10.1016/j.copsyc.2021.09.023

Singler JV. The sociohistorical context of Liberian English. Journal of Pidgin and Creole Languages. 2006;21(2):241–57.

Skutnabb-Kangas, T. (1981). Bilingualism or not. Multilingual Matters.

Skutnabb-Kangas, T. (1984). Bilingualism or not: The education of minorities. Multilingual Matters.

Smith, M. E. (1939). Some light on the problem of bilingualism as found from a study of the progress in mastery of English among pre-school children of nowAmerican ancestry in Hawaii. Genetic Psychology Monographs, 21, 119–2

Stevens, J. P. (2012). Applied multivariate statistics for the social sciences (5th ed.). Routledge.

Sui, X., Massar, K., Kessels, L. T. E., Reddy, P. S., Ruiter, R. A. C., & Sanders-Phillips, K. (2021). Violence exposure in South African adolescents: Differential and cumulative effects on psychological functioning. Journal of Interpersonal Violence, 36(9–10), 4084–4110. 10.1177/0886260518774184

Sylvestre, A., Bussières, È.-L., & Bouchard, C. (2016). Language problems among abused and neglected children: A meta-analytic review. Child Maltreatment, 21(1), 47–58. 10.1177/1077559515616703

Tabachnick, B. G., & Fidell, L. S. (2019). Using multivariate statistics (7th ed.). Pearson. https://www.pearsonhighered.com/assets/preface/0/1/3/4/0134790545.pdf

Tajfel, H., & Turner, J. C. (1979). An integrative theory of intergroup conflict. In W. G. Austin, & S. Worchel (Eds.), The social psychology of intergroup relations (pp. 33–37). Monterey, CA: Brooks/Cole.

Thabet, A., Al Ghamdi, H., Abdulla, T., Elhelou, M. and Vostanis, P. (2010) Attention deficit-hyperactivity symptoms among Palestinian children. Eastern Mediterranean Health Journal, 16, 259–263.

Tran, V., Verdon, S., McLeod, S., & Wang, C. (2022). Family language policies of Vietnamese–Australian families. Journal of Child Science, 12(1), e67–e78. 10.1055/s-0042-1743490

Trent, M., Dooley, D. G., & Dougé, J.; Section on Adolescent Health, Council on Community Pediatrics, & Committee on Adolescence (2019). The impact of racism on child and adolescent health. Pediatrics, 144(2), e20191765. 10.1542/peds.2019-1765 (publications.aap.org)

Turay M. Can English serve as the national language of Sierra Leone? Res J Engl Lang Lit. 2019;7(4).

Uktamovna, R. Z. (2025). The Transformative Journey of Adolescence: A Study of the Physical, Cognitive, Emotional, and Social Changes During the Teenage Years. Spanish Journal of Innovation and Integrity, 39, 169–172. Retrieved from https://sjii.es/index.php/journal/article/view/270

UNESCO. Education in a Multilingual World. Paris: UNESCO; 2003. (Education Position Paper).

United Nations Department of Economic and Social Affairs. (2024). The sustainable development goals report 2024. United Nations. https://unstats.un.org/sdgs/report/2024/

United Nations Educational, Scientific and Cultural Organization. (2025). Multilingual education: Global guidance on multilingual education and inclusion. https://www.unesco.org/en/languages-education/need-kno

United Nations General Assembly. (1951). Convention relating to the Status of Refugees, 189 U.N.T.S. 137. Article 1A(2). https://www.unhcr.org/3b66c2aa10

United Nations High Commissioner for Refugees. (2021). Persons at risk. UNHCR Emergency Handbook. https://emergency.unhcr.org/protection/persons-risk

United Nations. The Sustainable Development Goals report 2024. New York (NY): United Nations; 2024.

United Nations. The Sustainable Development Goals report 2025. New York (NY): United Nations; 2025.

USCR. USCR country report Sierra Leone: statistics on refugees and other uprooted people, Jun 2001 [Internet]. ReliefWeb; 2001 Jun 19 [cited 2025 Sep 21]. Available from: https://reliefweb.int/report/guinea/uscr-country-report-sierra-leone-statistics-refugees-and-other-uprooted-people-jun

van der Kolk BA. (2003).The neurobiology of childhood trauma and abuse. Child Adolesc Psychiatr Clin N Am. 2003;12(2):293–317, ix. doi: 10.1016/s1056-4993(03)00003-8.

van Tubergen, F. (2010). Determinants of second language proficiency among refugees in the Netherlands. Social Forces, 89(2), 515–534. 10.1353/sof.2010.0092

Veltman, C. (1983). Anglicization in the United States: Language environment and language practice of American adolescents. International Journal of the Sociology of Language, 44, 99–114. 10.1515/ijsl.1983.44.99

Vlasenko, V. V., Rogers, E. G., & Waugh, C. E. (2021). Affect labelling increases the intensity of positive emotions. Cognition and Emotion, 35(7), 1350–1364. 10.1080/0269

Vygotsky, L. S. (1978). Mind in society: The development of higher psychological processes. Harvard University Press.

Warren, A. (2025). Loneliness as a driver of allostatic load: Mechanisms linking social disconnection to physiological dysregulation and health disparities. Stress, 28(1), Article 2594067. 10.1080/10253890.2025.2594067

Weldemariam, K. (2022). The home literacy environment as a venue for fostering bilingualism and biliteracy: The case of an Ethio-Norwegian bilingual family in Oslo, Norway. Journal of Early Childhood Literacy, 22(4), 603–626. 10.1177/14687984211062398

Yağmur, K., & van de Vijver, F. J. R. (2012). Acculturation and language orientations of Turkish immigrants in Australia, France, Germany, and the Netherlands. Journal of Cross-Cultural Psychology, 43, 1110–1130. 10.1177/0022022111420145

Yarseah A, Adegoroye AOS. The experience and dimensions of shame as predictors of PTSD among Liberian refugees in Nigeria. Int J Res Anal Rev. 2019;6(2):1–8.

Yarseah DA, Ogunsanmi OO, Ogunsanmi JO, Ibimiluyi OF, Olaoye EO, Ezeani ES, Cheeseman VH. The mediating effects of perceived social support and shame on psychological distress and its dimensions among Liberian refugees in Nigeria. PLOS Ment Health. 2025;–. doi:10.1371/journal.pmen.0000330

Yeomans-Maldonado, G., & Mesa, C. (2021). The association of the home literacy environment and parental reading beliefs with oral language growth. Reading Research Quarterly, 56(4), 701–716. 10.1002/rrq.384

Zárate-Alva, N. E., & Sala-Roca, J. (2019). Socio-emotional skills of girls and young mothers in foster care. Children and Youth Services Review, 100, 50–56. 10.1016/j.childyouth.2019.02.036

